# Method of combining multiple researches to determine non-infectious disease causes. Analysis of Depression and Celiac disease causes

**DOI:** 10.1101/2023.12.24.23300513

**Authors:** Alan Olan

## Abstract

In this work an author is introducing a method which using a special algorithm based in math, allows to find disease causes for a specific non-infectious disease using results of multiple researches regarding risk factors of the disease. The method is based on a model presented in the article *“A Connection between Factors Causing Diseases and Diseases Frequencies: Its Application in Finding Disease Causes” (Alan Olan,* Journal of Clinical Trials, Vol.13, Issue 4.) which is confirmed by empirical data. As explained in the article mentioned above, a non-infectious disease is caused by a combination of few physiological parameters changes beyond 1-sigma interval which are existing at the same time. In the current article author explains a foundation of the method and then shows a practical way to use it. Using this method author further analyses Depression and Celiac diseases and as a result the work gives the causes of these diseases as a set of *physiological parameters changes beyond 1-sigma* interval and also as a set of disease causing external factors which combination in an individual *must c*ause these diseases as per presented model. Using the method an author shows why Depression has 2 causes and Celiac Disease has 6 causes. Author’s introduction to the method will allow other medical researchers to use their own and existing researches to determine the causes of non-infectious diseases as per presented model, using a simple algorithm.

## Introduction

In article [1] *“A Connection between Factors Causing Diseases and Diseases Frequencies: Its Application in Finding Disease Causes” (Alan Olan*, Journal of Clinical Trials, Vol.13, Issue 4.) we introduced a model of non-infectious disease. According to this model which is matching to empirical evidence, the non-infectious disease is caused by changes to multiple physiological parameters when their values go beyond 1-sigma interval, slightly less actually. This means a non-infectious disease must occur if 2 or more particular physiological parameters changes beyond 1-sigma exist for while at the same time. Also, based on the model the criteria was introduced to determine if a risk factor is causing a non-infectious disease or not. In order to be a cause of non-infectious disease the risk factor **K** calculated as (RR - 1) or (OR-1) to be **3.55+/-50%** (or 355%+/-50%) if the factor is causing a change in **1** physiological parameter, and should have value of **19.67+/-50%** (1967%+/-50%) if it is causing a change to 2 physiological parameters beyond 1-sigma.We call these conditions a **Disease Causation criteria.** The Disease Causation criteria determines whether a factor is one of multiple which are causing disease. The disease causing factor cannot cause a disease as a standalone as per model in article [1] because it is usually changing only 1 physiological parameter out of multiple required. Non-infectious disease, as per the model presented in article [1], is caused by multiple physiological parameters changed beyond 1-sigma.

The article [1] introduced a formula to calculate number of non-infectious disease causes if the disease rate (incidence rate usually) is known for the disease in a specific population. The formula is shown below (**n** is number of disease causes (as number of physiological parameter changed beyond 1-sigma) :

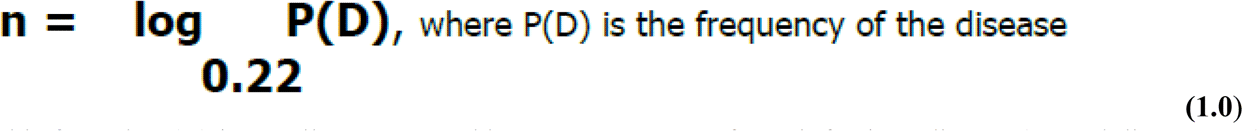

In this formula P(D) is usually represented by **incidence rate** of non-infectious disease (annual disease rate). Using formula 1.0 and Disease Causation criteria it was shown how to find physiological parameters changes which are causing some non-infectious diseases. In this article we introduce a method which is based on this model, which allow researchers to analyze an existing research about a specific non-infectious disease and determine *a full set* of disease causing physiological parameters for a non-infectious disease in much more complicated cases. Also, a method would allow to map a found disease causing factor to a physiological parameter which is changed by the factor beyond 1-sigma interval. For example, if we know that Diabetes is a disease causing factor for Hypertension there is a need to know which specific physiological factor impacted by Diabetes is really a cause of Hypertension. There are multiple physiological factors impacted by Diabetes and the method will allow to find a single one out of so many (allows finding “a needle in a haystack”) which is really causing Hypertension.

The method we introduce is based in math and those who mathematically inclined can find its foundation in the chapters of this article below. Here we will introduce a basic idea of the method and steps on how to practically use it.

## Explanation of the method

A method is introducing an *algorithm* based in math (but *not requiring* to use it much) which allows to process data from results of existing medical researches in few steps and *produce a new information* which consist of disease causes represented by a set of **physiological parameters changes beyond 1-sigma** (slightly less, actually) and also, a few **separate groups of disease causing external factors**. These few groups of factors are such that if you take a disease causing factor from each group as a standalone it will *not* cause a disease but combined together with factors from all other groups *must* cause a non-infectious disease.

Let’s say using the formula for a number of Disease causes (1.0) we found the number of causes as physiological parameter changes beyond 1-sigma. We also found from an existing research a few factors which are really causing a specific disease using a Disease Causation criteria provided. Now we need to determine which specific physiological parameters are really causing a disease. We only know their number and don’t know yet what they are. We know the factors which are causing a change to these physiological parameters and we can use them to find these unknown parameters.

In order to do this we need to list many physiological parameters related to the factor and we can find them from existing scientific research (for example, we can look for all biomarkers of Diabetes and list as many as we can find). Once we list the physilogical parameters in the header of some table we need to list the factors found to be causing the disease along a most left column (vertically) of the same table. Based on the researches’ data we already know which physiological parameters are related to those factors causing disease and we mark the intersection between the factor and the related parameter with letter “R” (or other letter). Please note, we use a term “related” as it is unknown if the physiological parameter change is a cause a disease or not yet and all we know is that its change is present if the factor is present. Now we start analyzing which factors are having same parameter in common and we will call it **an *intersection* if unrelated disease causing factors A and B both having the same physiological parameter changed.** We need to find all these intersections. Those intersections will be a superset of the physiological parameters which are causing a disease. The details of why this happens are provided in a detailed math explanation in this article. Here we can say that a match in just one physiological parameter changed (*important*: we looking for **a match by name**, not by value) between 2 different factors is not a random coincidence but a pattern. This pattern is determined by a fact that factors are causing a change of the physiological parameter beyond 1-sigma interval (or sometimes by other reasons explained later).

Here is *a simplified* explanation how the method works. Suppose there is a non-infectious disease with 2 unknown disease causes (quantity of 2 we have determined using formula 1.0). Suppose we determined already 2-3 disease causing factors (we can do it using Disease Causation criteria).Each disease causing factor is changing physiological parameters randomly beyond *their original(or normal) range* which means a quantity of physiological parameters changes is random, the **parameters’ names**, values are different as well, etc. The physiological parameters changes for a disease causing factors are random as they cannot be determined unless we do an experiment and determine what they have changed under influence of each of these disease causing factors (we assume we do an experiment first time). Note, the physiological parameter changes are *random for a disease causing factor* but they may be almost *the same for different people* impacted by this factor.

Suppose via the experiments we found that a disease causing factor 1 has changed 20 different physiological parameters beyond their original (or normal) range and a disease causing factor 2 has changed 50 different physiological parameters beyond their original (or normal) range. We just explained that *only 2 unknown* physiological parameters are really causing this disease. In order to determine **their names** we use a fact that due to random “selection” of physiological parameters out of so many (few thousands and more exist in human body) by ***unrelated*** *disease causing* factors, a match by **their names** and values between these 2 groups of physiological parameters practically is not possible by accident (this is shown mathematically in this article). If we find **a match of physiological parameters by name it is a pattern** under these specified conditions, it is not random. For example, if one type of *disease causing* factors (suppose a chocolate consumption) is causing a change in hemoglobin levels and a totally different *disease causing* factor (suppose a long exercising) is causing a change in hemoglobin levels as well then it is a pattern *under these specified conditions*.It becomes a pattern which we are interested in *because* it happens between groups belonging to *2 different disease causing* factors. We know these 2 *disease causing factors* are impacting same physiological parameter, meaning a parameter with the **same name**. For this reason, in order to find a pattern (in our case a disease causation physiological parameter) we need to find a place where 2 physiological parameters changes in both groups are the **same by name** (*important*: not by value) for these 2 different disease causing factors. This place where 2 disease causing factors have **the same name** of physiological parameter changed beyond original(or normal range) we call “an intersection”. If we found such a match between physiological parameters **by name** (*important:* not by value) we found a pattern - a physiological parameter which is causing a disease. See a pic 1. Also, in Appendix 1 we provide a simple analogy which could help to clarify or visualize the explained concept further.

**Pic 1.**
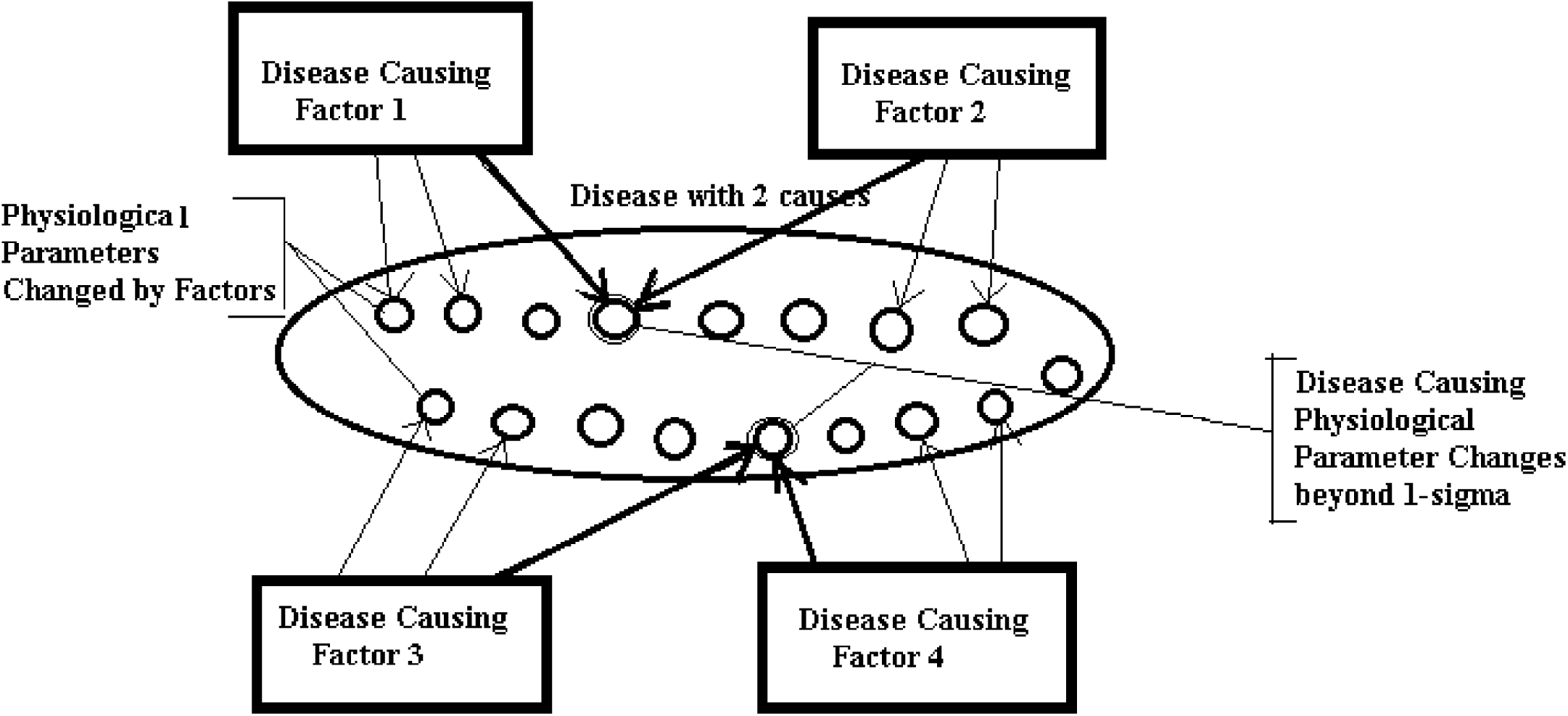
(Disease with 2 causes, as per formula (1.0): Disease causing factors are causing multiple physiological changes. Disease causing physiological changes are common for Factors 1 and 2, for Factors 3 and 4. We find them by determining an intersection in a physiological parameter)

When we find all these intersections we will find a list (or a matrix) of the parameters which are superset of physiological parameters which changes go beyond 1-sigma and are causing a disease. As there might be other patterns causing a match in parameters the number of parameters can exceed the number of parameters we determined via Number of Disease Causes formula (1.0). We will need to eliminate other parameters based on the methods provided in this article to get a final set of Physiological Parameters which changes go beyond 1-sigma (actually slightly less).These changes when they are present at the same time *must* trigger a disease. The illustration of this process shown on Pic 1.

Once we know the final set of these physiological parameters and as we know up front which factor is related to which physiological parameter we can find which specific physiological parameter is impacted by the specific factor in such a way that a parameter goes beyond 1-sigma interval. In other words we get an understanding that a factor is impacting a specific physiological parameter. Now, if an individual is affected by a set of factors which are causing a change in ALL these physiological parameters required then the individual will get sick after sometime unless the harmful factors are removed fast enough. This knowledge can help to prevent a disease in the individual or help potentially cure it or reduce severity of the disease.

## Using a method in practice

In a nutshell, in order to use this method to find a set of physiological parameters which are causing a disease next steps should be taken:

1. Find the number of causes (as physiological parameters changes) for the non-infectious disease using a formula for a disease causes *(1.0) (see also our article:, **“*****A Connection between Factors Causing Diseases and Diseases Frequencies: Its Application in Finding Disease Causes”**, *Alan Olan*, Journal of Clinical Trials, Vol.13, Issue 4.)
2. Find as many as possible, the factors which are causing a disease using a Disease Causation criteria on existing experimental data or new research (*see our article: **“*****A Connection between Factors Causing Diseases and Diseases Frequencies: Its Application in Finding Disease Causes”**, *Alan Olan*, Journal of Clinical Trials, Vol.13, Issue 4.)
3. For each factor found, you need to list as many as possible parameters related to it in some table. For example, if a factor is a consumption of some food then include which nutrients, chemicals it contains including harmful ones as per existing researches. If a factor is a disease then list which physiological parameters are known to be impacted one way or another as a symptom, a cause, etc. You need to place one factor vertically and appropriate impacted physiological parameters horizontally (in the table’s header) in this table. The column where a factor is related to a physiological parameter can be marked with cross “X” or letter “R”.
4. If the factor is by itself a physiological parameter it can be taken as one of the physiological parameters you are searching for during this step in the method. It can be corrected with next step if needed.
5. Now, you need to find where the columns which contain physiological parameters are causing a crossing between factors. This means you need to **look for a cross (“X” sign) in the same column for 2 or more factors in the table**. We are basically finding which same (by name, not value) physiological parameter is changed by 2 disease causing factors.
6. Build a list or a matrix **Pm** consisting of the parameters which are causing instersections. For example, like Pm: { r1, r19, r23, r45 }. In practice Pm can look like **Pm = { “Blood Pressure”, “Estrogen Level Change”, “Oxygen level”, “ROS level” }**
7. Eliminate redundant physiological parameters from matrix **Pm** if required (as per rules provided in this article). If the number of calculated parameters per formula (1.0) is less than number of parameters found via analysis of intersections then the redundant parameters need to be eliminated. The number of parameters found via analysis of intersection in a table should match to the number calculated via a formula (1.0) or be less if not all parameters can be found due to a lack of experimental data, etc. Also, if some physiological parameters found as result of analysis of intersection in a table are causing some disease causing factors to change more parameters then predicted by disease causing criteria then some physiological parameters may need to be eliminated as well. The disease causing factor can usually change only 1 or 2 physiological parameters beyond 1-sigma interval (slightly less than this interval, actually).
8. Verify the results of using algorithm by checking if there are appropriate experimental data consistent with the physiological parameters found. Also, you can verify if a physiological parameter found satisfies a Disease Causation criteria above ***if*** there is any research on the risk of this parameter.

You can see that **a method allows to use a simple algorithm** on a set of existing experimental data in order to determine disease causes (as a set of physiological parameter changes beyond 1-sigma interval) and **then verify the results using other research** (which was not used doing steps of the algorithm) either by using Disease Causation criteria or by simply checking how this research is consistent with results of your findings.

## Example of method’s application for a disease with 2 causes

Let’s look at hypothetical disease with 2 causes and apply to it the method described above. We created a Table 1. We can see in **Table 1** that all physiological parameters are listed on the top and all found from the experiments Factors which are causing this hypothetical disease are listed on the left. Now, we can see that parameter **r20** is an intersection for Factor 3 and Factor 4, param **r12** is an intersection for Factor 2 and Factor 3 and Factor 4. A param **r17** is an intersection for Factor 1, Factor 2. We can build our matrix **Pm : { r12, r20, r17}**. In practice, this matrix may look like a list **Pm : { ‘Blood Pressure Change’, ‘Oxygen Level Change’, ‘Estrogen Change’).** This matrix contains physiological parameters which changes beyond 1-sigma are causing a disease if present at the same time.

**Table 1.** (Areas in gray are where an intersections happen in parameters r12, r20 and r17 accordingly. Area surrounded by bold frame is a final set of parameters for matrix Pm)

| **Factors causing Disease** | **Physiological Parameters - Entire Set for ALL FACTORS as R1, R2, ... Rn** |  |  |  |  |  |  |  |  |  |  |  |  |  |
| --- | --- | --- | --- | --- | --- | --- | --- | --- | --- | --- | --- | --- | --- | --- |
|  | **r12** | **r20** | **r2** | **r8** | **r17** | **r75** | **r34** | **r56** | **r39** | **r12** | **r55** | **r23** | **r78** | **r11** |
| **Factor 1** |  |  | x |  | x |  |  |  | x | x |  | x |  |  |
| **Factor 2** | x |  |  |  | x |  |  | x |  |  | x |  |  | x |
| **Factor 3** | x | x |  |  |  |  | x |  |  |  |  |  |  |  |
| **Factor 4** | x | x |  |  |  | x |  |  |  |  |  |  | x |  |

Now, we see the matrix **Pm : { r12, r20, r17 }** has 3 elements but our disease only require 2. We need to eliminate the incorrect one. We can notice that if we take a valid combination of 2 param as {r12, r17 } then a param r12 is showing that *Factor 2 is impacting r12 and r17 parameters but Factor 2 only can cause 1 parameter* impact (as determined by Disease Causation criteria) so the combination is not correct. If we eliminate r12 and take a combination of {r20, r17 } then both params are satisfying our requirement (assuming there is no info their changes are within 1-sigma). So from this table we can see that final physiological parameter’s set which is causing this disease is **{r20. r17 }**.In this case Factors 3 and 4 are causing a change in parameters **r20**, Factor 1 and 2 are causing a change in physiological parameter **r17**. We has determined 2 physiological causes for a disease with 2 causes.

## Mathematical foundation of the method

Here we will go into a mathematical foundation of the method in details.Let’s look at the simple case of a disease where it was prior determined via experiments that multiple factors are causing changes in **only 2** physiological parameters of human body (parameters further).

Let the factors be **F1, F2, … Fn** and the parameters be **C1, C2.** Now, let’s look at the case where *F1, F2,…, Fn factors separately causing **only 1** change* in physiological parameters either C1 or C2 beyond 1-sigma (this actually often takes place in practice). That means that only C1 *or* C2 changes by some factor Fj (j E {1,2,..n }). Let’s **P1, …, Pn**, where n > 2 be *the sets of all physiological parameters which are related to factors F1, F2, .. Fn* accordingly. For example, P1: { r12, r15, r29, 43 }, P2: { r15, r28, r34, r89, r34, r12, r98 }, etc.

Let’s look at standalone factor **Fj.** As we know, a factor **Fj** (where *j is some integer from 1 to n*) impacts the specific physiological parameters either **C1 or C2** then we know that *this params C1 or C2 should be part of its set of* **Pj** as **it contains ALL the related to Fj parameters (a complete set) .** *Let’s take a factor F1 such that its set P1 contains **C1**, and choose some F2 such that its set P2 contains **C2*** (it is possible as we know factors impact either C1 or C2*), then if we choose any other factor as F3 then its set of P3 must contain either C1 or C2 (as F3 also impacts these physiological parameters - either C1 or C2 and P3 is a complete set). If P3 contains C1 then it intersect with P1. If P3 contains a C2 it intersects with P2. So **P3 must intersect with either P1 or P2 (either in C1 or C2).** In similar way we can apply this to P4, P5,… Pn. So this brings us to conclusion that **a set of physiological parameters Pn, where n > 2 must intersect with either P1 or P2 either in C1 or C2.** This means parameters Pn intersect with each other either in C1 or C2.* We can see representation of this set’s behavior on the Pic 2. ***All sets Pj, Pk,… matching to Fj, Fk,.. on the Pic 1 are crossing and only in C1 or C2 but not both.***

**Pic 2.**
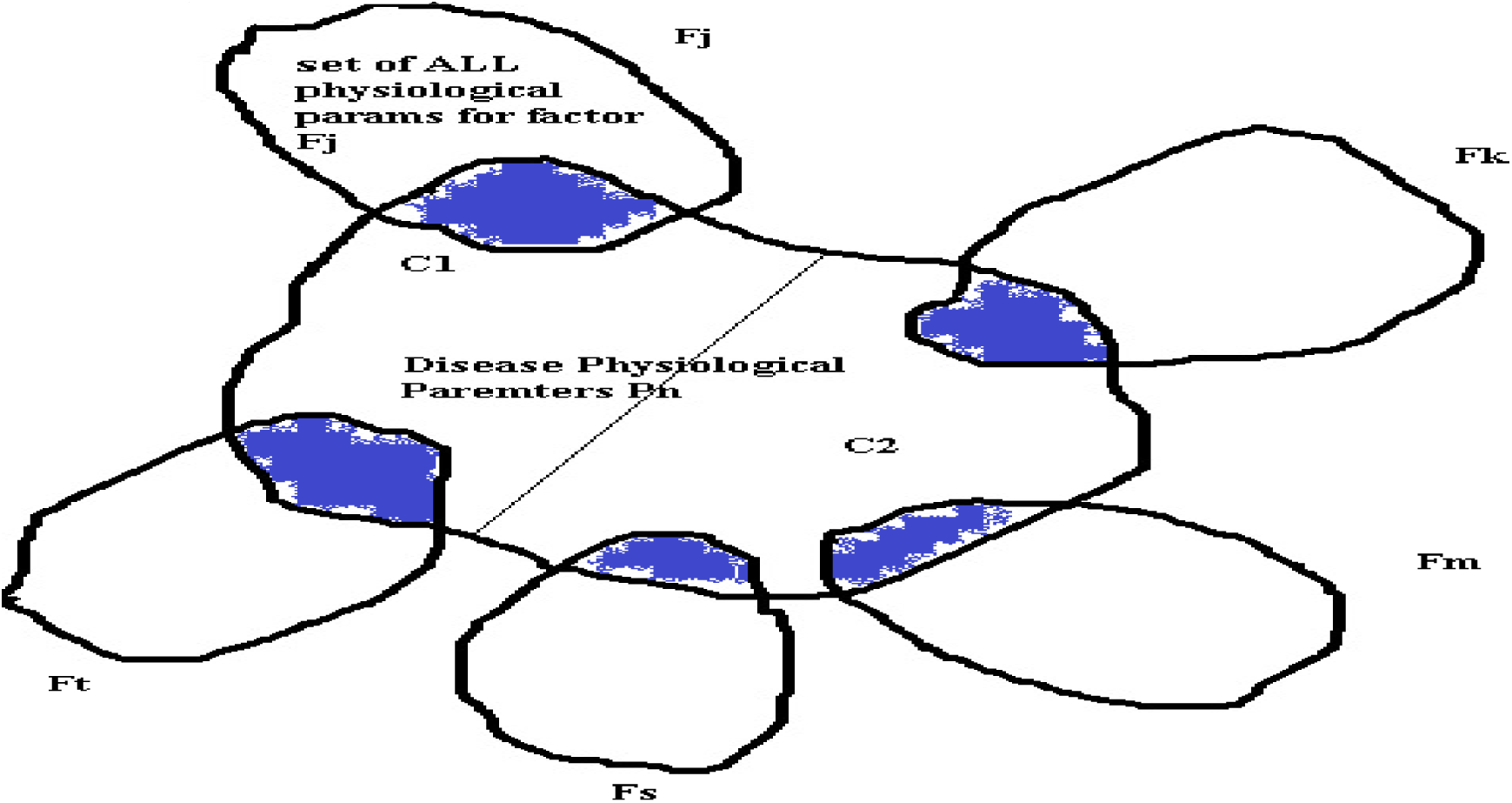
(Blue areas are area where Pj, Pk, etc for factors Fj, Fk, etc. intersect with a set of physiological parameters C1, C2 which are part of set Pn and which are causing diease)

We don’t know the values of C1 and C2 but if *we can find where parameters Pn intersects with each other we can determine **a subset** of physiological parameters **Pm**:* { Ry, Rx, Rz,.., Rt } which contains values of C1 and C2. This subset of **Pm** will be much smaller then set of all possible params included in P1, P2, .. Pn (as it is a subset and similarities in params of P1,.. Pn are not very probable and that is addressed below) but may contain more then 2 parameters and *only 2 parameters of this subset **Pm*** can be real physiological parameters causing a disease as they are C1 and C2.

In order to eliminate the incorrect parameters from subset **Pm** we need to notice:

1. that the params C1 or C2 should ***be such so all P1, P2, …, Pn intersect in them*** and if some parameters of ***Pm***: { Ry, Rx, Rz,.. Rt } don’t fit this rule *their need to be eliminated.* Practically it means this. We take random (or using a common sense) a combination of some 2 parameters **Rk** and **Rm** from a set of **Pm** *and check if the P1, … P2 all intersect in them if not then the Rk and Rm combination is not a valid set of C1, C2 and we may need to check another set of 2 parameters **Rk** and **Rg***
2. if some parameter of set ***Pm***: { Ry, Rx, Rz,.. Rt } ***is known as not changed beyond 1-sigma it should be eliminated*** as disease is caused by change in param beyond 1-sigma (as per our model).
3. if some parameter of ***Pm***: { Ry, Rx, Rz,.. Rt } ***is causing some set Pn intersect 2 times with some other set Pk then it should be eliminated*** as factors F1, F2, .. Fn can *only* impact 1 physiological parameter in this case and cannot impact / intersect 2 or more due to this.

The method above was described for a case of factors F1, …, Fn impacting ***only 2*** parameters but it can be extended to 3 and more parameters.

## How likely are random matches between physiological parameters?

As we discussed above the set of physiological parameters ***Pm***: { Ry, Rx, Rz,.., Rt } where we observe intersections may contain more parameters then needed (more than 2 in our case and due to other reasons). We need to be concerned with a question such as if we find one intersection of sets P1 and P2 in a physiological parameter belonging to 2 different external factors how likely it can be a random intersection? To answer this question let’s formulate the problem mathematically.

Let’s have a set A of integers from k = 1 to very large N. Let’s randomly select **n** numbers in set of P1 = {Ak, Ag, .., At } and then randomly select **n** numbers into set P2 = { Af, As, … Al } from our original set A (k= 1 to N) such that each element repeats only once in set P1 and only once in P2 (it is a unique element to sets P1, P2). For example, if we chose a number 3 as part of the set P1 then it only exist one time in the set P1. What is a probability that we find element **Ai** in set P1 and P2 ?

To anwer this question let’s do next steps. Let’s limit set A by some top element enumerated by **t** (so set is not infinite).

1. We can take **n** elements from **t** elements of set A with number ways **tCn**
2. Number of ways to take **n** elements with an element **Ai** equals the number of ways to select **n-1** elements (we exclude Ai) from **t -1** (set of A elements) and is **t-1Cn-1**
3. Then probability to take **n** elements which include element **Ai** in set P1 (or P2) is P(Ai E Psel) = **t-1Cn-1 / tCn,** where Psel is P1 or P2 sets
4. The probablity that element Ai will be in P1 and P2 is P (Ai E P1 and Ai E P2) = P (Ai E P1) * P (Ai E P2) as events independent.
5. So probability P (Ai E P1 and Ai E P2) = P (Ai E P1) * P (Ai E P2) = (**t-1Cn-1 / tCn) ^ 2**
6. Or finally, the probablity that element Ai wil be in P1 and P2 is P (Ai E P1 and Ai E P2) = (**t-1Cn-1 / tCn**) ^ 2

Using a formula above let’s calculate a probability of match in element Ai if we take randomly elements from a sequence of numbers from 1 to 1000 (t = 1000, assuming so many physiological parameters exist) and take only n = 10 element into sets P1 and P2 accordinly. P(Ai E P1) = **_999_C_9_** / **1000C_10_** = (2.63 * 10^21) /(2.63 * 10^23) = 1 / 10^2 = 0.01 the same is fare for P(Ai E P2) = 0.01 and so the probability of getting element Ai in sets P1 and P2 is

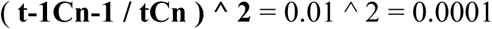

This is a probability of radom match. **The probability of non-random match** is **1 - P (Ai E P1 and Ai E P2)** = 1 - 0.0001 = 0.9999 ∼= 1 so very close to 1. It means *if we see a match between set P1 and set P2 in some element Ai it exremely likely it is not random.* This is an important conclusion. *The only matches we find practically are not random but are caused by some reason* and in our case it is due to same physiological parameter impacted by 2 different factors. We need to notice that number of paramaters actaully much more as most agree that there are around 20,000 different proteins in our body and each is a potential physiological parameter. So the probability of the match selecting 10 of 20,000 will be much smaller!

In practice there are about over **t=150** physiological parameters known to medicine and for a single factor we usually find about **n=30** related paramters. Doing calculations for this case we ge**t** P(Ai E P1) = **_149_C_29_** / **_150_C_30_** = (6.43 * 10^30) /(3.2 * 10^31) = 0,2 and so the probability of getting element Ai in sets P1 and P2 is

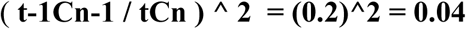

We see the probability of random match is higher in practice (4%) and so in practice we can see more random matches. The probability of at least one random match will also increases as we find intersections for dozens of different causation factors.

This random matches still can be eliminated with methods described in this article by applying other restrictive conditions, including using our criteria for disease causes.

## Criteria for choosing intersections of physiological parameters

As we could see there is a very small chance of random intersection of physiological parameters belonging to 2 different factors and also, there are cases when more parameters found then determined by disease causation criteria. In order to select those parameters which are really causing a disease we need to use these criteria below.

1. We need to check if factors which are impacting physiological parameters found to be intersections are belonging to **same type of disease** or they are **very similar in nature** (like BMI impact and high weight impact) and we can ignore them as the intersection is most likely due to similarity.
2. Make sure **a factor is causing a change the physiological parameter** which intersect (not opposite) As the factors should cause a change in physiological parameters..
3. **Make sure both factors are causing change in the physiological parameter which intersect** or the intersection is not valid.
4. If a **factor causing insignificant change (less 1-sigma interval) in physiological parameter then the parameters need to be ignored as a point of intersection** with this factor. The factors as we discussed need to make change in physiological parameter so its value will be beyond *1-sigma* interval.
5. Make sure the **physiological parameters changes in the point of intersection happen in the same direction** (either increase or decrease). If the changes move in different directions then the parameters are considered as 2 different parameters and not as one.
6. **The intersection of parameters for a similar factor is not considered correct** intersection (as it is the same factor, for example Increased Weigh and BMI over 30 are similar factors)
7. If the experiment found that a factor which causing a disease is impacting 1 physiological parameter but the factor consists of combinations of 2 separate factors (for example observation was done for presence of diabetes or obesity together so they work as a factor) then **the combined factor can be allowed to have 2 intersections despite having impact on 1 physiological parameter** as one of the parts of the factor could be present at the time (for example either a diabetes causing 1 change in physiological parameter or obesity causing the change in another physiological parameter)
8. **Element cannot cross itself**, for example if factor is IgA and physiological parameter IgA then they don’t create a crossing. Usually, if a factor found as physiological parameter then we already know this is a physiological parameter causing a disease.
9. In many cases it is possible to **eliminate the redundant intersection by determining** via a known research **if the physiological parameter increases or decreases with the presence of specific factor.** For example, does lipoprotein level decreases or increases with diabetes as causation factor? If **same parameter where we see a intersection, increases in presence of one factor and decreases in presence of another** then the intersection is invalid.
10. Once a set of physiological parameters was found **they can be also validated with already provided Disease Causation criteria to determine if the factor is a cause of disease** (a parameter should increase a risk of disease 3.5 +/-50% times) using an existing research. For example, if we find that an increased lipoprotein is causing a stroke and if some existing research confirms that a risk of its increase is **3.5 +/-50%** then it confirms a lipoprotein as a parameter was found correctly.
11. In some cases biochemical analysis may help to eliminate an element from matrix Pm

## METHODOLOGY

### Analysis of Non-Infectious Disease Causes

Below we provide analysis of causes of 2 non-infectious diseases - Depression and Celiac disease as per method presented above. Using these 2 examples we demonstrate in more details how to use a method at 1^st^ on relatively simple analysis case of Depression and then on a more complex case of analysis of Celiac disease which requires elimination of redundant physiological parameters. We also provide an approximate algorithm of doing parameters elimination.

#### 1. Depression

Depressive disorder (also known as depression) is a common mental disorder. It involves a depressed mood or loss of pleasure or interest in activities for long periods of time. It causes severe symptoms that affect how a person feels, thinks, and handles daily activities, such as sleeping, eating, or working.

Major depression is affecting more than 8% of American adults each year (https://mhanational.org/conditions/depression#:~:text=Basic%20Facts%20About%20Depression,are#20affected%20by%20major%20depression.). Calculating number of causes using formula (1.0) we get:

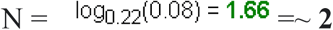

As per calculations above the depression is caused by changes to **2** physiological parameters beyond 1-sigma.

Let’s list the factors which are causing a Depression and explain how they are determined based on Disease Causation criteria provided above and developed in article ***“*A Connection between Factors Causing Diseases and Diseases Frequencies: Its Application in Finding Disease Causes”** *(Alan Olan,* Journal of Clinical Trials, Vol.13, Issue 4.) :

1. Study shows: “Working 11+ hours a day was related to a 2.43-fold odds of MDE compared to working 7 to 8 hours a day in an analysis adjusted for socio-demographic characteristics”, “It was also robust to additional adjustment for work characteristics (job strain and social support at work), **the odds ratio of MDE being 2.52-fold for 11+ working hours in the final mode**l.” (*“Overtime Work as a Predictor of Major Depressive Episode: A 5-Year Follow-Up of the Whitehall II Study*”, Marianna Virtanen, Stephen A. Stansfeld, et al, Plos One, published: January 25, 2012, https://doi.org/10.1371/journal.pone.0030719). We get **OR = 2.52 which is the range of 4.5+/-50%(2.27-6.81)** and it means the factor is one of 2 causes of Depression and impact 1 physiological parameter (as per Disease Causation criteria). We list this factor in Table 1.1 (Factors column)
2. Another study about Major Depression Disorder (MDD) shows: “individuals with insomnia but without, any psychiatric disorders were also more likely to develop a new-onset MDD in the subsequent year (**OR=5.4,**95% CI=2.6-11.3) compared with individuals with neither insomnia or psychiatric disorders.”, “In a longitudinal study of 979 young adults,<u>^24^</u> **insomnia increased the relative risk for depression fourfold** (95% 0=2.2-7.0) over a 3-year period” (*“Sleep disturbances and depression: risk relationships for subsequent depression and therapeutic implications”*,Peter L. Franzen, PhD*, Dialogues Clin Neurosci. 2008 Dec; 10(4): 473–481, doi: 10.31887/DCNS.2008.10.4/plfranzen)) . As we can see **OR = 4** is **the range of 4.5+/-50%(2.27-6.81)** and **OR =5.4** as well, so it means insomnia is a causation factor for a depression and is one of 2 causes of depression and is impacting 1 physiological parameter. We list this factor in Table 1.1 (Factors column)
3. Study shows: “In addition, initial presentation with **social phobia was associated with** a **5.7-fold increased risk of developing major depressive disorder**.” (‘The Critical Relationship Between Anxiety and Depression”, Ned H. Kalin, M.D., The American Journal of Psychiatry, Published Online:1 May 2020 https://doi.org/10.1176/appi.ajp.2020.20030305). If **OR =5.7** it is within range of **4.5+/-50%(2.27-6.81)** of Disease Causation criteria for 1 physiological parameter. It means social phobia is a causation factor for major depressive disorder and impacting 1 physiological parameter. We list this factor in Table 1.1 (Factors column)
4. Study shows: “Subgroup studies indicated that **epilepsy was associated with an increased risk of depression in Asian, African and Caucasian populations** (Asian: **OR/RR = 2.42**; 95% CI: 1.48-3.95; **African: OR/RR = 2.48**; 95% CI: 1.88-3.28; Caucasian: OR/RR = 1.86; 95% CI: 1.60-2.15). Subgroup studies showed that **epilepsy was associated with an increased risk of depression among adolescents and adults** (adolescents: **OR/RR = 2.54**; 95% CI: 1.86-3.46; adults: OR/RR = 2.22; 95% CI: 1.79-2.75)” (*“Association between epilepsy and risk of depression: A meta-analysis”,* Chu Chu, Psychiatry Research, Volume 312, June 2022, 114531). We can see that **OR = 2.42**, **OR/RR = 2.48, OR/RR = 2.54** are within a Disease Causation criteria **4.5+/-50%(2.27-6.81) .** It means that Epilepsy is one of the causes of depression and impacts 1 physiological parameter. We list this factor in Table 1.1 (Factors column)
5. Study shows that: **“women in late transition to menopause were nearly 3 times more likely to report these depressive symptoms compared with pre-menopausal women** (**OR, 2.89**; 95% confidence interval [CI],1.29-6.45; *P* = .01).” (*“Hormones and Menopausal Status as Predictors of Depression in Womenin Transition to Menopause”*, Ellen W. Freeman, PhD; Mary D. Sammel, ScD; Li Liu, MD, MS and et al, Arch Gen Psychiatry. 2004;61(1):62-70. doi:10.1001/archpsyc.61.1.62). We can see that **OR = 2.89** and it is in within a range of **4.5+/-50%(2.27-6.81)** as per Disease Causing criteria. It means that being a women in late transition to menopause is a disease causing factor which impacts 1 physiological parameter out of 2 which are causing depression. We list this factor in Table 1.1 (Factors column).
6. Research on Seasonal affective disorder (seasonal depression) .”An analysis of sequence variations in three genes that form a functional unit of the circadian clock (**PER2, ARNTL, and NPAS2**) found a **single nucleotide polymorphism** in each gene with a significant association with seasonal affective disorder. The authors also reported an additive effect and identified a risk genotype combination and a protective genotype combination. **They reported that carriers of a risk genotype have a four-fold risk of having seasonal affective disorder** than other genotype combinations and a ten-fold risk compared to those with the protective genotype”(*“Seasonal affective disorder (seasonal depression) – symptoms, risk factors, causes and genetics”*, Medicover Genetics, Medicover Genetics Editorial Team, November 16, 2022). If we take four-fold risk as **OR= 4** then it is within a range for Disease Causation criteria **4.5+/-50%(2.27-6.81).**It means a particular PER2, ARNTL, and NPAS2 **single nucleotide polymorphism** is a causation factor for a depression and impacting 1 physiological parameter. We list this factor in Table 1.1 (Factors column).

We know now some of the factors (which we could find from research, there may be others) which are causing Depression but we need to find out: **1)** which physiological parameters are really changed by this factors to cause a disease. We know there are 2 physiological parameters like this via a calculation using formula (1) above but they names are unknown yet. **2)** we need to find whether the causation factors we found impact 1st or 2nd of these physiological parameters. We want to find out for each factor which physiological parameter (1st or 2nd) are impacted by this specific factor. See Pic 2 for illustration.

**Pic 2.**
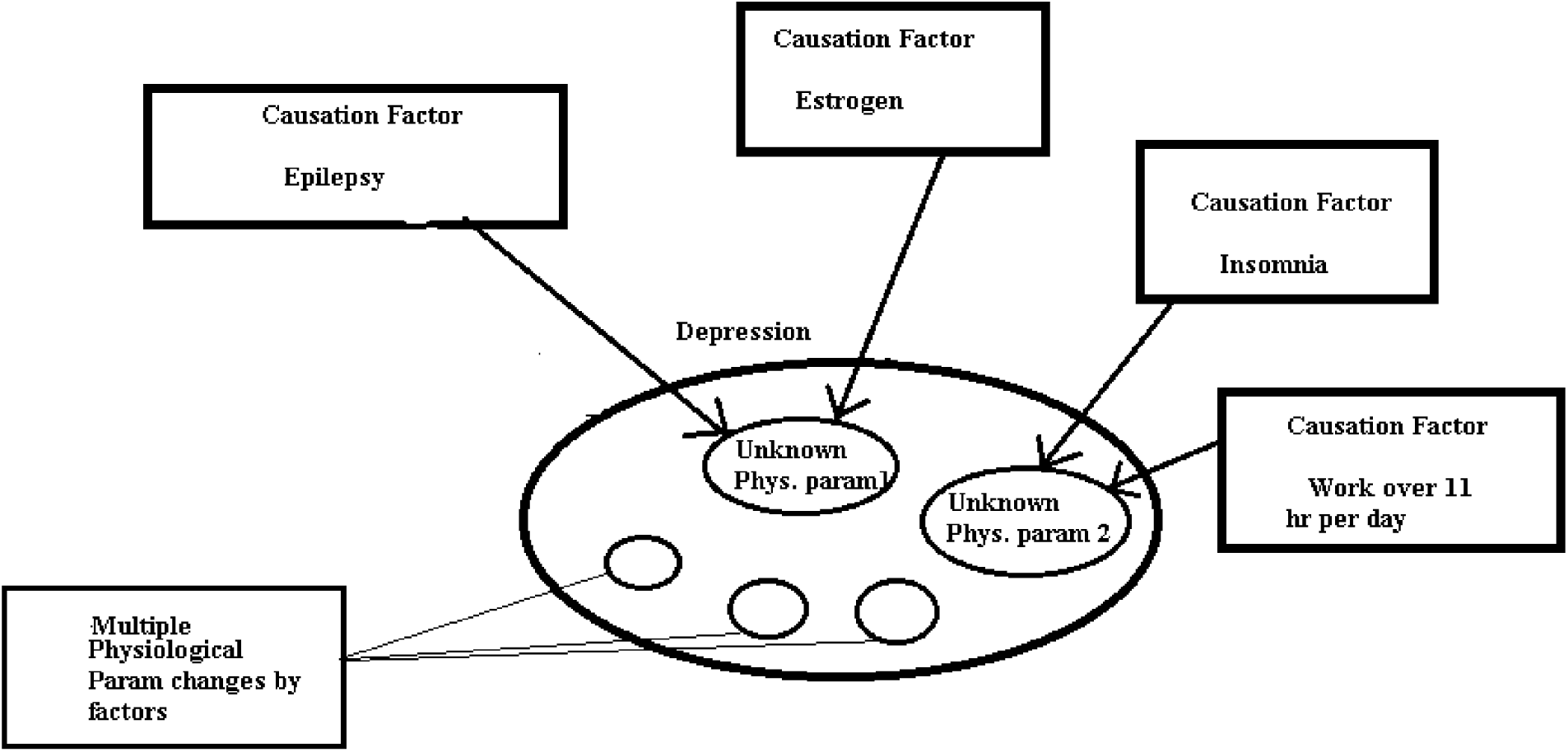
(Illustrates how different causation factors are impacting same 2 disease causing physiological changes in Depression. Notice how different factors “intersect” in the same physiological parameter)

In order to determine this we have listed the factors above in a Table 1.1 vertically (*as an alternate way*) in factor’s column (1) and will find as many as possible the physiological parameters which are related to these factors and list them horizontally(in the table’s header). We mark the relationship between a factor and a physiological parameter in the table with letter “R”. It means there is a change to a physiological parameter when the factor is present. As we can see one factor may be related to changes to multiple physiological parameters. We need to find out the intersections of the parameters across the factors as explained in the method above. These intersections, after elimination of invalid parameters (if they exist) will give us a set of 2 physiological parameters changes beyond 1-sigma which **must** cause a Depression when present at the same time. We stress **these physiological parameters changes beyond 1-sigma must cause the Depression if co-exist, it is not optional** as per the model presented in article [1].

Here we provide a list of physiological parameters related to factors of depression as citations from respective studies(note: our bold font):

1. “Two studies showed **increased oxygen consumption across 24 h in insomnia patients** compared with good-sleeping controls.”(“Is Metabolic Rate Increased in Insomnia Disorder? A Systematic Review”, Julia L. Chapman, Maria Comas, Camilla M. Hoyos, et al,Front Endocrinol (Lausanne). 2018; 9: 374, published online 2018 Jul 16. doi: 10.3389/fendo.2018.00374). Based on this we put this parameter in Table 1.1 as “High Oxygen consumption” in column (2) and add “R” letter for Insomnia there.
2. BDNF and Sleep Disorder: “Studies conducted up to this point on the subject of changes in **peripheral BDNF level in insomnia consistently show BDNF reduction in individuals afflicted with this sleep disorder**, regardless of sex or accompanying psychiatric conditions;” (*“Investigating the Role of BDNF in Insomnia: Current Insights”*, Ditmer M, Gabryelska A, Turkiewicz S, Sochal M, Nature and Science of Sleep, 7 December 2023 Volume 2023:15 Pages 1045—1060). Based on this we put this parameter in Table 1.1 as “Low BDNF” in column (5) and place letter “R” for Insomnia
3. Social phobia in animal studies: “According to an experimental study with laboratory animals, **stress by social subjugation is able to foster hyperalgesia with a decrease in the BDNF levels;** this condition is more evident in “susceptible” subjects rather than in “resilient” ones” (*“BDNF Protein and Anxiety Disorders”*, Tatiana Marins Farias, Rebeca Ataíde Cerqueira, Danton Ferraz Sousa, et al, Neurological and Mental Disorders, Published: 28 May 2020). Based on this we put letter “R” in Table 1.1 under column (5) for Social Phobia factor as related to it.
4. Long hour and burn out: “…study reported that **the serum BDNF (sBDNF) levels were significantly lower in burnout subjects than those in healthy people**. Moreover, sBDNF levels were negatively correlated with EE and CY, but positively with PE (Sertoz et al., 2008). Our **recent study also showed an inverse association between job burnout and sBDNF level in doctors and nurses** (He et al., 2017). “(*“Interaction between job stress, serum BDNF level and the BDNF rs2049046 polymorphism in job burnout”*,Shu-Chang He, Shuang Wu, Chao Wang, Dong-Mei Wang et al, Journal of Affective Disorders, Volume 266, 1 April 2020, Pages 671-677). Based on this we put letter “R” in Table 1.1 under column (5) for a factor Working long hours (1 cs).
5. “Mechanistically, single-particle tracking revealed **that E2 [Estradiol] treatment selectively reduced the dwell time and thereby decreased the confinement of GABAA Rs** at inhibitory synapses.”and also “In summary, **our studies provide a molecular mechanism by which estrogen acts to reduce the efficacy of GABAergic inhibition** by decreasing the stability of inhibitory synapses.” (“Estradiol modulates the efficacy of synaptic inhibition by decreasing the dwell time of GABAA receptors at inhibitory synapses”, Jayanta Mukherjee, Ross A. Cardarelli, Yasmine Cantaut-Belarif, et al, PNAS, Vol. 114 | No. 44, https://doi.org/10.1073/pnas.1705075114). Based on this we put this physiological parameter in Table 1.1, column (4) and put a letter “R” for a factor “Women in late transition to menopause (high estrogen)” as women have significant level of estrogen comparing to men and its levels change periodically.
6. “the present study reveal that **estrogen decreases the GABA_B_ receptor-mediated autoinhibition of GABAergic** POA neurons. In addition, **estrogen induces an apparent decrease in the subsequent ability of these neurons to synthesize GABA**.” (“Estrogen Biphasically Modifies Hypothalamic GABAergic Function Concomitantly with Negative and Positive Control of Luteinizing Hormone Release”, Edward J. Wagner, Oline K. Rønnekleiv, Martha A. Bosch, and Martin J. Kelly, J Neurosci. 2001 Mar 15; 21(6): 2085–2093, doi: 10.1523/JNEUROSCI.21-06-02085.2001). Based on this we put “**Estrogen Modulation of GABA**” in Table 1.1, column (3) and mark the factor of “Women in late transition to menopause (high estrogen)” with letter “R” under this column.
7. Role GABA in Epilepsy: “Experimental and clinical study evidence indicates that **GABA has an important role in the mechanism and treatment of epilepsy**: (a) Abnormalities of GABAergic function have been observed in genetic and acquired animal models of epilepsy; (b) **Reductions of GABA-mediated inhibition**, activity of glutamate decarboxylase, binding to GABAA and benzodiazepine sites, GABA in cerebrospinal fluid and brain tissue, and GABA detected during microdialysis **studies have been reported in studies of human epileptic brain tissue**; “ (*“GABAergic mechanisms in epilepsy”*, D M Treiman, Epilepsia 2001:42 Suppl 3:8-12. doi: 10.1046/j.1528-1157.2001.042suppl.3008.x.). Based on this we put a letter “R” under “Reduction of GABAergic inhibition” in Table 1.1, column (4) and place a letter “R” in Epilepsy under this column.
8. Role GABA in Epilepsy: “**Reductions of GABA mediated inhibition** and decreased activity of GAD **has been reported in studies of human epileptic brain tissue**“ (*“Decreased GABA receptor in the cerebral cortex of epileptic rats: effect of Bacopa monnieri and Bacoside-A”*, Jobin Mathew, Savitha Balakrishnan, Sherin Antony, Pretty Mary Abraham & CS Paulose, Journal of Biomedical Science, 19, Article number: 25 (2012)). We already place a letter “R” under **“Reduction of GABAergic inhibition”** column for Epilepsy.This is 2^nd^ confirmation of relationship between elilepsy and GABA mediated inhibition.

Let’s analyze and find these intersections of the physiological parameters for the factors impacting Depression in Table 1.1. We checking in which column parameters intersect. We can see that only parameters in columns 4 and 5 are causing intersection for all set of factors listed. We mark the intersection in gray color to differentiate them.Remember, we are looking for physiological parameters match by name (not by value).

The areas which are gray out in Table 1.1 show the intersections of physiological parameters but they are not yet a final list. Only once we eliminate redundant parameters (if they exist) then we can surround the final parameters with bold rectangles.

By looking at the intersections of physiological parameters in Table 1.1 we can write down a matrix Pm = {**“Reduction of GABAergic inhibition”**, **“Low BDNF”** } which has only 2 physiological parameters so we don’t need to eliminate any. We also observe that this matrix satisfies a hard condition that factors should only intersect in 1 physiological parameter. This matrix represent a set of 2 physiological parameters changes beyond 1-sigma which if present simultaneously will trigger depression within some time. **We can list them like below:**

1. Reduction of GABAergic inhibition beyond ∼1 sigma interval
2. Low BDNF (beyond ∼1 sigma)

The **columns in Table 1.1 surrounded by bold** rectangles represent the physiological parameters found as result of our analysis and which are causing a disease.

There is a factor of Circadian clock genes **PER2**, ARNTL, and **NPAS2** SNP which should be causing impact to 1^st^ or 2^nd^ physiological parameters of the Depression. Which one is it? Is this **“Reduction of GABAergic inhibition” or “Low BDNF”?** As this factor is causing only 1 impact to physiological parameter then it should be impacting only one of them to be beyond 1-sigma. The existing research below suggests that these Circadian clock genes’ single nucleotide polymorphism(SNP) should cause **“Reduction of GABAergic inhibition”:**

1. One research informs**: “**These results: (1) implicate Npas2 in the response to stress and the development of anxiety; and (2) provide **functional evidence for the regulation of GABAergic neurotransmission by NPAS2 in the ventral striatum** (*“NPAS2 regulation of anxiety-like behavior and GABAA receptors”*, Ryan Wellington Logan,Frontiers in Molecular Neuroscience, November 2017)
2. Another research states: “Overall, our results provide evidence for a specific **role of glial Per2 in mood-related behavior, accompanied by dysregulation of components of the glutamatergic, GABAergic** and dopaminergic **signaling “** (*“Deletion of the clock gene Period2 (Per2) in glial cells alters mood-related behavior in mice”,* Scientific Reports, Tomaz Martini,1 Jürgen A. Ripperger,1 Jimmy Stalin,et al)
3. Another research states**: “The release of neurotransmitters**, such as dopamine, glutamate, and γ-**amino-butyric acid (GABA) have been shown to be modulated by circadian rhythms** (Castaneda et al., 2004). **Per2 is associated with the generation of the circadian rhythms** (Arjona and Sarkar, 2006; Sujino et al., 2007)(*”Neurobiological Functions of the Period Circadian Clock 2 Gene, Per2”*, Mikyung Kim, June Bryan de la Peña, et al, Biomol Ther (Seoul). 2018 Jul; 26(4): 358–367.)

On this example of determining the Circadian genes role in Depression you can see that even if we don’t know exact impact of the causation factor on a specific physiological parameter **using results of analysis** we still could determine just 2 possible options for this impact and we are able to suggest which option will likely be correct one based on existing research. Based on this we can place letter “R”as suggested for the “Circadian clock genes **PER2**, ARNTL, and **NPAS2** SNP” factor in Table 1.1 in column 4 (“**Reduction of GABAergic inhibition”)** to signify the relationship between the genes’ SNP and reduction of GABAergic inhibition,

Now let’s take a look at empirical evidence related to the found physiologic causes of Depression by looking at existing researches which are provided below:

1. One article informs that: “Studies have demonstrated **low concentrations of γ-aminobutyric acid (GABA) in the plasma and CSF of individuals with major depression,** and low GABA concentrations have also been found in the occipital cortex of depressed subjects***“*** *(“Increased Occipital Cortex GABA Concentrations in Depressed Patients After Therapy With Selective Serotonin Reuptake Inhibitors”* (by Gerard Sanacora, M.D., Ph.D., Graeme F. Mason, et al, The American Journal of Psychiatry, 1 Apr 2002) and it is consistent with a reduction of GABAergic inhibition found in analysis according to our method. Another article provides a similar view that *“*Evidence suggesting that central nervous system γ-**aminobutyric acid (GABA) concentrations are reduced in patients with major depressive disorder (MDD)** has been present since at least 1980, and this idea has recently gained support from more recent magnetic resonance spectroscopy data. These observations have led to **the assumption that MDD’s underlying etiology is tied to an overall reduction in GABA-mediated inhibitory neurotransmission**.***“****(“Altered γ-aminobutyric acid neurotransmission in major depressive disorder: a critical review of the supporting evidence and the influence of serotonergic antidepressants*“ (by Pehrson A, Sanchez C, Drug Design, Development and Therapy, published 19 January 2015 Volume 2015:9). This also consistent with analysis a per our method which has found that a reduction of GABAergic inhibition is a disease cause for Depression by *using not directly related data - from researches on epilepsy and insomnia!*
2. An article states: “**The role of BDNF in depression has gained broad attention because many pre-clinical and clinical studies provide direct evidence** suggesting that modulation in **expression of BDNF** could be involved in behavioral phenomenon associated with depression.”(*“Brain-derived neurotrophic factor: role in depression and suicide”* (by Yogesh Dwivedi, Neuropsychiatr Dis Treat. 2009; 5: 433–449.) which is also consistant with analysis’s result made according to our method that a deficiency of neurprotective peptides (BDNF) is one of the causes of depression. Another article informs : “The **studies mentioned in this review article greatly support the role of BDNF in the pathogenesis of depression** and treatment of this disorder with antidepressants.”(*“Unfolding the Role of BDNF as a Biomarker for Treatment of Depression”* (by Tarapati Rana, Tapan Behl, et al, Journal of Molecular Neuroscience November 2020, volume 71, pages2008–2021 (2021)) and also consistant with BDNF as disease cause of depression found algorithmicaly via our method with a use of mutliple available researches on factors **not directly related to BDNF** (the research we used was related to insomnia, social phobia and long working hours)!

We can see that existing research generally in agreement with result of our method, the research points to Reductions of GABA mediated inhibition and BDNF levels. Important to notice here that conclusions per our method were done based on researches on risk factors which **were not related to GABA or BDNF**. Using our method we came to these conclusions by using a research for **the factors which were causing disease** and they were not *explicitly referring* to Reductions of GABA mediated inhibition or BDNA level. Our results came from analysis of factors causing disease by using a pretty simple algorithm but these results gave a totally *new set of knowledge* of disease causation for Depression disease and which is consistent with an empirical evidence.

Now let’s recap what we done. We calculated number of causes for Depression based on the incidence rate and determined a number of causes as 2. Then we found the factors which are causing depression using a Disease Causation criteria for a Risk factor (or OR) related to 1 physiological parameter change beyond 1-sigma. We listed the found factors in the Table 1.1 along with related physiological parameters. By looking at the table we found intersections where physiological parameters are same *by name* for different factors. Based on these intersections we built a matrix **Pm** with physiological parameters. We analyzed the **Pm** matrix and found there is no redundancy in parameters as we found 2 and this is expected to be 2. So **we found the physiological parameters causing disease purely from knowing disease causing factors by the simple algorithm** and we can see that **they match to the experimental data**! It shows the algorithm works.

Based on the analysis of external factors **a depression is caused by simultaneous co-existence of 2 physiological parameters changes beyond ∼1 sigma:**

1. Reduction of GABAergic inhibition beyond ∼1 sigma interval
2. Low BDNF (beyond ∼1 sigma)

**It means** a person will have a depression disease if his/her GABAergic inhibition decrease beyond ∼1 level AND there will be a low level of BDNF (beyond ∼1 sigma).

Now we can move data from **Table 1.1 to Table 1.2** which is a table of results and can be used for results validation by checking number of cases(physiological parameters changes) found for a specific factor with quantity determined by a Disease Causation criteria for the factor.

Now we can move data from the Table 1.2 to Table 1.3 which will show the 2 physiological parameters causing a Depression and also show which factors are impacting them. By looking at this table we can determine which external factors will cause Depression in an individual if combined together with another one

**Table 1.1.** **R** - means that the physiological parameter is related to the factor **cs** - a cause (means a factor is causing a physiological parameter change beyond 1-sigma, 1cs - 1 parameter changed)

| **Factors causing Depression** | **Physiological Parameters for Factors ( ALL SET) causing Depression** |  |  |  |
| --- | --- | --- | --- | --- |
|  | **High Oxygen Consumption** | **Estrogen Modulation of GABA** | **Reduction of GABAergic inhibition** | **Low BDNF** |
| **(1)** | **(2)** | **(3)** | **(4)** | **(5)** |
| Insomnia (1cs) | **R** |  |  | **R** |
| Social Phobia (1 cs) |  |  |  | **R** |
| Women in late transition to menopause (high estrogen) (1 cs) |  | **R** | **R** |  |
| Epilepsy (1 cs) |  |  | **R** |  |
| Circadian clock genes PER2, ARNTL, and NPAS2 SNP (1 cs) |  |  | R (suggested) |  |
| Working long hours (1 cs) |  |  |  | **R** |

**Table 1.2.** For example, if we take Epilepsy from the column 1 and combine it with Insomnia from column 2 then we can determine that once an individual has this combination he/she should develop a Depression at some point of time unless one of these factors eliminated.

| **External Factors** | **Physiological parameters which changes are causing Depression** |  |
| --- | --- | --- |
|  | **Reduction of GABAergic inhibition** | **Low BDNF** |
| **(1)** | **(2)** | **(3)** |
| Insomnia (1 cs) |  | **x** |
| Circadian clock genes PER2, ARNTL, and NPAS2 SNP (1 cs) | **x**<br>(suggested ) |  |
| Epilepsy(1 cs) | **x** |  |
| Working over 11 hour daily(1 cs) |  | **x** |
| Social phobia (1 cs) |  | **x** |
| Women in late transition to menopause (high estrogen) (1 cs) | **x** |  |

**Table 1.3.**
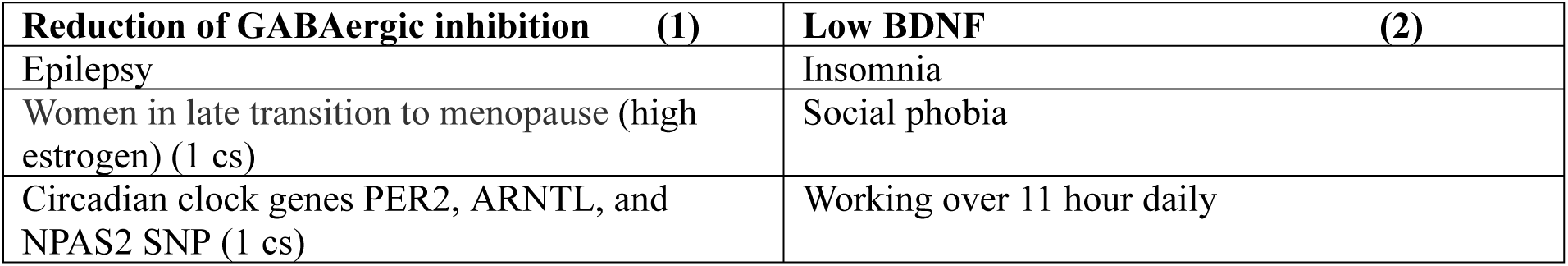
External factors which can cause a depression shown in Table 1.3. Any factor *combination out of 2 different* columns should cause depression.

| **Reduction of GABAergic inhibition (1)** | **Low BDNF (2)** |
| --- | --- |
| Epilepsy | Insomnia |
| Women in late transition to menopause (high estrogen) (1 cs) | Social phobia |
| Circadian clock genes PER2, ARNTL, and NPAS2 SNP (1 cs) | Working over 11 hour daily |

**Note:** the list of factors in table 1.3 is not complete and based on research known to an author. There can be other disease causing factors but they still should impact these 2 physiological parameters as these parameters are really causing a disease. (There might be few exceptions to this rule though as some factors may have caused a change to a frequency of the disease over the time.)

### Building a hypothesis of disease pathology knowing physiological parameters causing the disease

As we could see we have found *a structure of mechanism of Depression disease* - we have determined that it consist of “Reduction of GABAergic inhibition beyond ∼1 sigma interval” and “Low BDNF (beyond ∼1 sigma)”.

But **knowing a structure of mechanism of the disease (the physiological parameters causing the disease) allows us to build a hypothesis of pathology** of the disease by “connecting the dots” of this structure. For example, we can build next hypothesis of Depression pathology.

**Hypothesis of Depression pathology** (using 2 found physiological parameters causing a disease): GABAergic inhibition is getting reduced beyond -1 sigma by actions of disease causing factors (epilepsy, estrogen changes, etc) and this should increase excitation of neurons which in sequence should increase a demand for BDNF. At the same time due to a low level of BDNF (beyond 1-sigma) already being present because of other disease causation factors (Insomnia, Working over 11 hours, etc) the increased demand to BDNF cannot be satisfied and this disrupts the normal work of the brain and cause a Depression.

As demonstrated here by knowing multiple physiological parameters causing a specific non-infectious disease determined using a proposed method it is getting more easy to build a hypothesis of the disease pathology.

The hypothesis then can be proved or rejected with future research.

#### 2. Celiac disease

Celiac disease is a serious auto immune disease that occurs in genetically predisposed people where the ingestion of gluten leads to damage in the small intestine.

The incidence of Celiac disease among women was 17.4 per 100,000 person, compared to 7.8 per 100,000 person among men. (https://celiac.org/about-the-foundation/featured-news/2020/02/incidence-of-celiac-disease-steadily-increasing/). Using a formula for number of disease causes let’s determine a number of physiological parameter changes which are causing Celiac Disease.

For women:

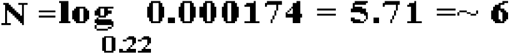

For men:

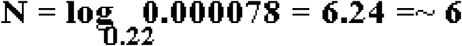

As per calculations for both incidence rates Celiac disease happens due to **6** physiological parameters’ changes.

Using a Disease Causation criteria provided in this article and developed in article ***“*A Connection between Factors Causing Diseases and Diseases Frequencies: Its Application in Finding Disease Causes”** (*Alan Olan*, Journal of Clinical Trials, Vol.13, Issue 4.) we are finding a list of factors based on existing research data which are really causing Celiac disease as they are changing some physiological parameter beyond 1-sigma. The list of found **<u>Celiac disease causing factors is below</u>** with related explanation for each factor:

1. “**having 3 or more respiratory infections in the first 6 months** of life **multiplied the risk by 2.8** (**OR 2.79**, 95% CI; 1.03–7.54). “ (*“Infections in early life as risk factor for coeliac disease”*, Sandra Llorente Pelayo, Mirian Palacios Sánchez, Pablo Docio Pérez, et al, Anales de Pediatría (English Edition), Volume 94, Issue 5, May 2021, Pages 293-300). As per our criteria for one physiological parameter impact the Risk Factor K should be **3.5** (and OR = K + 1 = 3.55 +1 = 4.55) and we have this OR = 2.8 which is within **4.55 +/- 50%** (2.27 - 6.81) and this means this factor related to infection is one of 6 causes of Celiac Disease. We place this factor in Table 2.1, column 2. Please note that we use an alternate way to populate a table here as we put the factors on the top of the table..
2. “**Intake of gluten before 2 years of age increases risk** of celiac disease at least 2-fold in children with genetic risk factors for this disease”, also “…children who received amounts of gluten in upper tertile (i.e. high gluten intake) were at more than **2-fold higher risk for CD** than those consumed less (**OR=2.65**; 95% CI=1.70–4.13; p <.0001(*“Effects of Gluten Intake on Risk of Celiac Disease: a case–control study on a Swedish birth cohort”*,Carin Andrén Aronsson, MSc, Hye-Seung Lee, PhD, Sibylle Koletzko, PhD, et al,Clin Gastroenterol Hepatol. 2016 Mar 14(3): 403–409, doi: 10.1016/j.cgh.2015.09.030). **OR = 2.65** is within **4.55 +/- 50%** (2.27 - 6.81) so this factor is a one of 6 causes of Celiac Disease as per criteria developed in article (***“*A Connection between Factors Causing Diseases and Diseases Frequencies: Its Application in Finding Disease Causes”**, *Alan Olan*, Journal of Clinical Trials, Vol.13, Issue 4.) . We place this factor in Table 2.1, column 3.
3. “Early clinical studies estimated a 10–**20 fold increased** risk for celiac disease (CD) **in IgA deficient subjects** [7], [8], [9]. This has recently been confirmed by several studies conducted on very large cohorts of IgA deficient patients [10], [11]” (*“Testing for IgG class antibodies in celiac disease patients with selective IgA deficiency.: A comparison of the diagnostic accuracy of 9 IgG anti-tissue transglutaminase, 1 IgG anti-gliadin and 1 IgG anti-deaminated gliadin peptide antibody assays”*, Danilo Villalta, Maria Grazia Alessio, Marilina Tampoia, et al, Clinica Chimica Acta, Volume 382, Issues 1–2, July 2007, Pages 95-4). According to a Disease Causation criteria in article [1] **the factor which is causing a disease and impacts 2 physiological parameters beyond 1-sigma should have a risk K = 19 +/- 50%** and so for **OR=20**, K = OR - 1 = 20 - 1 = 19 is within this range and it means the IgA related factor is one of the 6 causes a Celiac Disease and it is changing 2 physiological parameters. We place this factor in Table 2.1, column 4.
4. “The HLA-DQ2*5 molecule is encoded by DQA1*05:01 and DQB1*02:01 alleles in cis-configuration in the DR3 haplotype. In general, up to 95% of patients with CD are positive to **HLA-DQ2** (DQA1*0501/DQB1*0201), and the remaining 5% are positive to HLA-DQ8 (HLA-DQB1*0302) haplotypes.31 **HLA-DQ2/DQ8 common haplotypes have been shown to elevate** disease risk **by 6-fold**. “ (*“Understanding Celiac Disease From Genetics to the Future Diagnostic Strategies”*,Carolina Salazar, Jennyfer M García-Cárdenas and et al, Clinical Medicine Insights: Gastroenterology, Volume 10, 2017). Taking **OR = 6** we determine **K = OR - 1 = 6 - 1 = 5** and it is within **3.5+/-50% range** and so the factor is one of 6 cause of Celiac Disease, impacting **1 physiological parameter** change. We place this factor in Table 2.1, column 5.
5. “**Use of iron supplements reported at 18 months of age** was associated **with a three fold higher risk of celiac**” and more specifically the research shows that for child’s use of dietary iron supplements at 6 and 18 months the risk of celiac disease **OR = 3.1**, 95 % CI (*“Maternal iron supplement intake during pregnancy and risk of celiac disease in children*”, Ketil Størdal, Margaretha Haugen, Anne Lise Brantsæter, et al, Clin Gastroenterol Hepatol. 2014 Apr; 12(4): 624–631.e2). According to the Disease Causation criteria **K = OR - 1 = 3.1 - 1 = 2.1 and is within 3.5+/- 50% range** for a factor which is causing a disease by changing one physiological parameter. This means that High Iron supplementation factor above is **one of 6 causes of Celiac Disease**. We need to remind that only if all 6 physiologial changes take place then this will trigger a Celiac Disease. One factor as stand alone should not trigger the disease. For more details please see the article *“A Connection between Factors Causing Diseases and Diseases Frequencies: Its Application in Finding Disease Causes” (Alan Olan,,* Journal of Clinical Trials, Vol.13, Issue 4). We place this factor in Table 2.1, column 6.
6. Another study says - “ Polybrominated diphenyl ether (PBDEs), perfluoroalkyl substances (PFASs), and p,p’-dichlorodiphenyldichloroethylene (DDE) and HLA-DQ genotype category were measured in blood serum and whole blood, respectively.” and its results are “After stratifying by sex, we found **higher odds of celiac disease in females with serum concentrations of DDE** (**OR = 13.0**, 95% CI = 1.54, 110), **PFOS (OR = 12.8**, 95% CI = 1.17, 141), **perfluorooctanoic acid** (**OR = 20.6**, 95% CI = 1.13, 375) and in males with serum **BDE153, a PBDE** congener (**OR = 2.28**, 95% CI = 1.01, 5.18).”(*“Persistent organic pollutant exposure and celiac disease: A pilot study”*, Abigail Gaylord, Leonardo Trasande, et al, Environmental Research, Volume 186, July 2020, 109439) so we can find that Risk Factor K = OR -1 will be **K= 12 for DDE**, **K = 11.8 for PFOS**, **K** = **19.6 for perfluorooctanoic acid** which are all **within a 19+/-50% range** and matching to a Disease Causation criteria which impact 2 physiological parameter changes. This means **<u>DDE</u>**<u>, **PFOS, perfluorooctanoic acid found in serum concentrations** in females are really one of the **6**</u> **<u>causes of Celiac Disease impacting 2 physiological parameters.</u>**For males, **OR = 2.28** is in range between **4.55 +/- 50%** (2.27 - 6.81)and it mean **BDE153** is one of 6 causes impacting 1 physiological parameter. We need to notice here **these compounds are not causing the Celiac Disease as stand alone**. Only if 2 physiological parameters changes which are impacted by them present at the same time with other 4 out of 6 required to cause a disease changes only then Celiac Disease will be triggered as per mechanism described in article *“A Connection between Factors Causing Diseases and Diseases Frequencies: Its Application in Finding Disease Causes”(Alan Olan,* Journal of Clinical Trials, Vol.13, Issue 4).We place this DDE, PFOS, perfluorooctanoic acid related factor in Table 2.1, column 7.
7. “A total of 11 studies with 1147 cases and 1774 controls were selected for this meta-analysis. The pooled **results indicated that TNF-α -308G>A polymorphism was associated with increased risk of celiac disease** (A vs G: OR=2.077, 95% CI=1.468-2.939, P=≤0.001; AA vs GG: OR=8.512, 95% CI=3.740-19.373, P=≤0.001; AA+AG vs GG: OR=1.869, 95% CI=1.161-3.008, P=0.010; **and AA+AG vs GG: OR=4.773**, 95% CI=3.181-7.162, P≤0.001” (“*ASSOCIATION OF TNF- α-308G>A POLYMORPHISM WITH SUSCEPTIBILITY TO CELIAC DISEASE: A SYSTEMATIC REVIEW AND META-ANALYSIS*”, Arq. Gastroenterol. 56 (01) Jan-Mar 2019 • https://doi.org/10.1590/S0004-2803.201900000-20). As we can see combination of AA+AG vs GG increase risk of Celiac disease with **OR = 4.77** which is within a range of OR to be a cause of disease - **4.55 +/- 50%** (2.27 - 6.81). It means this combination is impacting 1 physiological parameter and in this case this is a TNF-Alfa and is one of 6 cause of Celiac Dieases. Additionally we can notice that according to an article “The frequency percentage of GG, AG and **AA** genotypes in TNF-α cytokines at -308A/G in Azeri population was 70.8%, **26.7%** and 2.5%, respectively”(*“TNF-α gene polymorphism in Iranian Azeri population”*, Mohammad Asgharzadeh, Manouchehr Fadaee, Hamed Ebrahimzadeh Leylabadlo, et al, Gene Reports, Volume 19, June 2020, https://doi.org/10.1016/j.genrep.2020.1006511) and we can see that probability of combination fo AG is 26.7 percent or about 0.27 and interesting to notice that it is the probability which matching to **TNF-Alfa properties being beyond 1-sigma** interval and as we know to cause the disease the physiological parameter value should be beyond 1-sigma interval, slightly less (see article *Alan Olan, “A Connection between Factors Causing Diseases and Diseases Frequencies: Its Application in Finding Disease Causes”).* It means that a probability to have **TNF-α -308G>A polymorphism** is 0.267 which is required for a parameter to be beyond 1-sigma and be a disease causing factor as well. We place this factor in Table 2.1, column 8.

Once we listed disease causing factors in Table 2.1 we are adding physiological parameters related to these factors and mark relationship between them and related disease causing factors with letter “R” in the table. For brevity, we don’t list here all but only few citations to the research which was used to populate these parameters but for more details how it is done you can refer to Depression section of this article as well.

1. “In a recent study, **an iron-fortified micronutrient powder provided to Kenyan infants ranging from 6 to 10 months of age** caused an increase of several taxa from *Enterobacteriaceae* family, especially the pathobiont *E*. *coli*, and **a decrease of *Bifidobacterium* in their intestine** [95]”, “One of the oldest studies back in 1985, showed that **infants given an iron-fortified cow’s milk preparation had lower *Bifidobacterium*** but higher counts of *Bacteroides* and *E. coli* than infants receiving an unfortified cow’s milk preparation [92]” (“*Gut Microbiota and Iron: The Crucial Actors in Health and Disease”*, Bahtiyar Yilmaz and Hai Li, Pharmaceuticals 2018, 11(4), 98; https://doi.org/10.3390/ph11040098). Based on this we add a Bifidobacterium as physilogical parameter change related to iron supplementation in Table 2.1 column **6** and put letter “R” to mark this relationship.
2. “**Homeostatic IgA (which also functions as a natural antibody) is induced in the intestinal mucosa by continuous stimulation with commensal bacteria**, and is also detectable in circulation. Indeed**, little IgA is detected in the intestinal secretions and sera of germ-free** and neonatal mice and its production is restored soon after the colonization of commensal bacteria (2) “(*“Regulation of IgA Production by Intestinal Dendritic Cells and Related Cells”*, Front. Immunol.,Hiroyuki Tezuka,Toshiaki Ohteki, 13 August 2019, Volume 10 - 2019, https://doi.org/10.3389/fimmu.2019.01891). Another paper informs: **“Immunoglobulin A (IgA) is involved in the maintenance of gut homeostasis. Although the oral administration of bifidobacteria increases the amount of fecal IgA”,** (*“Enhancement of IgA production by membrane vesicles derived from Bifidobacterium longum subsp. infantis”*, Atsushi Kurata, Shino Yamasaki-Yashiki, Tomoya Imai, et al Bioscience, Biotechnology, and Biochemistry, Volume 87, Issue 1, January 2023, Pages 119–128, https://doi.org/10.1093/bbb/zbac172). We see there is a relationship between commensal bacteria such as bifidobacteria and IgA. We mark this relationship with letter “R” in Column 4 of Table 2.1 for bifidobacteria.
3. An article informs: “**The presence/absence of gluten in the diet can change the diversity and proportions of the microbial communities constituting the gut microbiota**. There is an intimate relation between gluten metabolism and celiac disease pathophysiology and gut microbiota; their interrelation defines intestinal health and homeostasis”, and also “The **main bacterial strains described as related to this gluten metabolism are Rothia, Staphylococcus epidermidis,** Streptococcus pneumoniae, Streptococcus mitis, **and Bifidobacterium. “**(*“Dietary Gluten as a Conditioning Factor of the Gut Microbiota in Celiac Disease”*, Karla A Bascuñán, Magdalena Araya, Leda Roncoroni, et al, Adv Nutr. 2020 Jan; 11(1): 160–174, doi: 10.1093/advances/nmz080). We can see that consumption of gluten is related to the **Rothia, Staphylococcus epidermidis, Bifidobacterium** and we mark this relationshipt in Table 2.1, column 3 for “Gluten under 2 year” with letters “R” for each of these bacteria. We mark it as there is dependency between these bacteria and gluten which impacts the bacterial diversity.
4. Article informs regarding Influenza A: “An important finding in this study is **that TNF-levels and IL-8 levels manifest delayed peaks** in the nasal lavage fluid. Thus, as alluded to above, **while TNF- is already significantly elevated on day 2 of infection** at the time of peak symptom scores, and TNF-administration can induce the systemic symptoms we observed “ (*“Local and Systemic Cytokine Responses during Experimental Human Influenza A Virus Infection”*, Frederick G. Hayden, R. Scott Fritz, Monica C. Lobo, et al, J Clin Invest. 1998 Feb 1; 101(3): 643–649.10.1172/JCI1355). This and other researches support the fact that infections increase TNF-alfa cytokine production. We mark this relationship for TNF -alfa and infections with letter “R” in Table 2.1, column 2..
5. “**Polymorphism in the TNF-α cytokine at position -308G/A affects the rate of production of this cytokine** so that the **AA genotype is associated with its high level.** The **GG genotype frequency was more abundant** in the present study than the other two genotypes, and because TNF-α is a pre-inflammatory cytokine, *its high level can be effective in susceptibility to inflammatory* and autoimmune diseases “ (*“TNF-α gene polymorphism in Iranian Azeri population.”*, Mohammad Asgharzadeh, Manouchehr Fadaee, Hamed Ebrahimzadeh Leylabadlo, et al, Gene Reports, Volume 19, June 2020, 100651). As per this study we see that AA genotype of TNF-α is associated with high level of this cytokine and we mark this relationship with letter “R” in Table 2.1, column 8.
6. An article on DDT and DDE pesticides states: “Compared to women with the lowest **plasma concentrations of DDT and DDE, those with the highest concentrations of both compounds had higher levels IL-1β, IL6, and TNF-α**.“ (“Exposure to DDT from Indoor Residual Spraying and biomarkers of inflammation among reproductive-aged women from South Africa”,Lea A. Cupul-Uicab, Riana Bornman, Janet I. Archer, et al, Environ Res. 2020 Dec; 191: 110088, doi: 10.1016/j.envres.2020.110088). This relationship between *DDE, etc and increased TNF-Alfa* we mark with “R” for TNF-Alfa in Table 2.1, column 7 (for a “**DDE, PFOS, perfluorooctanoic acid pesticide**“ factor).

We can see how the relationships between external factors causing Celiac disease and physiological parameters which are *related* to these factors are found and marked in the Table 2.1. In similar way we complete the rest of the Table 2.1 population.

As we can see the Table 2.1 lists the external factors on the top of the table which are causing Celiac disease and shows how they connected to the changes in specific physiological parameters (which are listed in the most left column).

Here and further below **cs** - stand for a cause. 1 cs, for example, means a factor is causing one physiological parameter change beyond 1 sigma interval. We use this abbreviation in Table 2.1.

**R -** means that an external factor is located in the column is related to the change in physiological parameter which located in the specific row of the table. In other words some factor “XYZ” is related to the change in physiological parameter located in the row of the table and marked with letter “R”.

Now we do a similar process as we did for Depression disease above but for Celiac disease it is more complicated as it has redundant physiological parameters which we need to be eliminated…Let’s find an intersection between physiological parameters related to the factors causing Celiac disease listed in the table in order to find the physiological parameters which are impacted by these factors and this way to determine the disease causing physiological parameters. The physiological parameters changes which we find are the real causes of Celiac disease . By looking at the table 2.1 we can see for example, that factors in columns 3,4,6 intersect as they all have a common physiological parameter change as **Bifidobacterium.** Factors in columns 2 and 3 also intersect as they have a common physiological parameter change as **IFN-Gamma,** etc. We marked this intersection in gray color to separate them better from others.

By looking at intersections in the Table 2.1 as explained we can build our matrix **Pm** which will at first contains all intersected physiological parameters and which will include next elements:

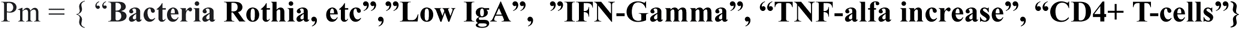

which contains 5 elements so at least 1 to be found yet. We include “Low IgA” in the matrix as it was determined already via Disease Causation criteria that it is a valid disease causing factor. Remember, the total number of physiological parameters causing Celiac was determined to be **6** using formula (1.0) in this article. Also, we need to check matrix **Pm** so *<u>all physiological parameters where intersection happens assure that each</u> <u>factor is impacting only predicted numbe</u>*<u>r *of physiological parameters*</u>, not more not less. For example, if a factor is impacting 1 parameter (we mark it as **1cs** in the Table 2.1 columns) then only one intersection for this factor should be observed. If we observe more intersections then some physiological parameters need to be eliminated from matrix Pm..

By looking at the table 2.1 we can notice that factor of “**Gltuten before 2 years (1 cs)”** is intersecting in **Bifidobacterium** and **IFN-Gamma, CD4+ T-cell** physiological parameters but it is not correct as this factors should be causing only 1 (per 1cs) physiological parameter change not 3. We need to resolve this issue by eliminating incorrect crossings.

We can do it this way. **“Low IgA**” factor should be crossing 2 physiological parameters (**2 cs**) and one of them IgA itself and another is **Bifidobacterium**. This condition satisfies if intersection happen in physiological parameters “IgA” and “Bacteria **Rothia, etc”.** In this case “IFN-Gamma” and “CD4+ T-cells” get excluded as Gluten can cross only in 1 parameter (1cs) and it was just determined to be **Bifidobacterium** and so cannot cross in these 2 parameters. So a matrix is now Pm = { “**Bacteria Rothia, etc”,”Low IgA”, “TNF-alfa**”}. Now we notice that “**Bacteria Rothia, etc”** is a group of bacteria, which we grouped here for brevity. Let’s take a look at this group. We can notice that factors in columns **3, 4,** 6 have intersection in **Bifidobacterium** and as there are 2 intersections it is a high priority pattern (it is highly unlikely, due to 2 intersections it is a random match).Factors in columns **3, 4** intersect 2 times (in bacteria Rothia and Staphylococcus epidermidis) and this can be ignored as intersection because a factor in column 3 and a factor in column 4 are already changing **Bifidobacterium** and were determined per Disease Causation criteria as factors changing only 1 physiological parameter (not more). Additional intersections in columns 3 and 4 *are patterns* too but it is likely that it happens due to relationship of dependent paramters (**Bifidobacterium** and these 2 bacteria). Based on this elimination we can write a final Pm matrix as

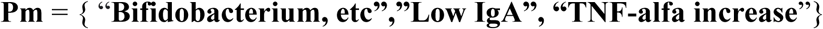

This matrix only has 3 physiological parameters now, physiological parameter number 4 is “Genetic Predisposition”(and 5, 6 are unknown yet) which was determined directly per disease causing criteria..

The question may arise why do we see 2 intersections between 2 factors in columns 2 and 3 which intersect in **CD4+ T-cell** and **TFN-Gamma**? As per our method a random intersection between 2 factors should be extremely rare and if it occurs it signifies a pattern with probability of at least 96%. But the pattern can be due to a fact that 2 parameters depend on each other, in this case a change **CD4+ T-cell** may indirectly cause a production of **TFN-Gamma** cytokine and by knowing the possible relationship of these factors one crossing can be potentially eliminated from matrix **Pm** as insignificant.

The **rows or columns in Table 2.1 surrounded by bold** rectangles represent the physiological parameters found as a final result of analysis and which are causing a disease.

### Algorithm to eliminate redundancy

An approximate algorithm which can be used to eliminate redundant physiological parameters may look like this:

1. Find out if all parameters which intersect in your table satisfy *a hard condition* that a factor only can cause as many changes to physiological parameters as determined via a disease causation criteria (which in most case is 1 and in some 2 or very rare 3).If some physiological parameters are causing not permitted number of changes by a disease causing factor then some parameters need to be eliminated (ignored as intersections).
2. Check the factors which having intersections. If the factors are similar or related then their intersections can be due to similarity and ignored.
3. Select a set of physiological parameters which from a common sense will make most of factors intersect in them. Validate if your selection is following a rule number 1) above. If not use the elimination criteria provided in this article to help resolve conflicts.
4. If 3 factors which are not related intersect in the same physiological parameter then treat a physiological parameter as a high priority intersection as it is extremely low probability that this intersection is not a pattern.
5. If a physiological parameter is selected as a real intersection surround it with a bold rectangle to differentiate from original intersections.
6. Remember that if 2 factors intersect it is a pattern which is determined by next facts 1) that factors are causing a same changes beyond 1-sigma to a physiological parameter **or** 2) the intersection occur due to dependency between related factors or related physiological parameters but the changes are not necessarily beyond 1-sigma. In other words in this case the intersection happens due to this relationships not necessarily due to 1-sigma change (but it can be both at the same time: due to relationship and also due to 1-sigma change)..

Now we create Table 2.2 for final results of analysis where we consolidate only the physiological parameter which cause the Celiac disease based on Pm matrix and add Genetic Predisposition. The table can be used to validate the results of analysis by checking a match that factors are causing as many changes to physiological parameters as it was determined by Disease causation criteria for the specific factor. For example, if a factor was determined to cause only 1 change in physiological parameter (**1 cs**) then we should observe this is the case in Table 2.2.

Now we can verify the results of analysis of Celiac disease completed above with existing research on Celiac disease below:

1. An article says “report on children during and after the Swedish celiac disease outbreak showed that rod-shaped bacteria constituted a significant fraction of the proximal small-intestine mucosal microbiota (<u>^94^</u>). The presence of these bacterial groups conferred a **4-fold increased risk** for celiac disease in children aged <2 y**”**(*“Dietary Gluten as a Conditioning Factor of the Gut Microbiota in Celiac Disease”*, Karla A Bascuñá, Magdalena Araya, et al., Advances in Nutrition, Volume 11, Issue 1, January 2020) . Based on a disease cause criteria it means **this bacterial disbalance is a one cause of the Celial disease as the risk is K= 4 -1 = 3 fold** (K = OR -1) and within **3.5+/- 50%** range.
2. Another research states “**The results of our study suggest a role for TNF-α in the pathogenesis of CD** [celiac disease - authors’ note] and demonstrate that T cells are a source of this proinflammatory cytokine. This finding is in keeping with reports that monoclonal antibody anti-TNF therapy has been employed successfully in a patient with refractory CD.”(*“TNF-α production by intraepithelial T cells in celiac disease”*, Conleth Feighery, Joan O’Keefe, Ph.D., Gastroenterology, VOLUME 125, ISSUE 5, P1560-1561, NOVEMBER 2003). This is consistent with our results of analysis that TNF-alfa change beyond 1-sigma is a physiological parameter which is causing Celiac disease if a change beyond 1-sigma.

As we can see the physiological parameters found to be cause of disease like bacterial change, TNF-Alfa increase as per method explained above are consistent with empirical data existing for Celiac disease causes. We also found that research on bacteria above suggests that bacterial change is one of the Celiac disease causes which consistent with the finding by our method.**We confirmed this result using a Disease Causation criteria** too.

We came to conclusions as per our method using *unrelated* to these physiological parameters research. We have used research on risk of gluten, respiratory infection and pesticides but *via a simple algorithm have produced new data* that bacteria changes(Bifidobacterium, etc) and TNF-alfa changes are causing Celiac disease which are consistent with related Celiac disease research.. It shows how that method works.

Overall according to the analysis done by a method explained above **Celiac disease is caused by simultaneous presence of these 6 physiological parameters changes beyond ∼1 sigma interval** but due to lack of experimental data we could only determine 4 of them:

1. Bifidobacterium, etc changes in the intestine beyond 1-sigma
2. Reduction of immunologlobulin IgA beyond 1-sigma
3. Genetic predisposition - HLA-DQ2/DQ8 common haplotypes
4. TNF-alfa increase beyond 1-sigma
5. **2 physiological parameters** beyond 1-sigma which need to be determined.

A Table 2.3 shows external factors which will cause a Celiac disease if they are taken from each column and combined together. A combination of 6 factors from each column will cause Celiac disease in an individual. If the factors represent the 2 causes (means they are changing 2 physiological parameters beyond 1-sigma) then the number of factors can be less accordingly.

**Table 2.1.** List of all physiological parameters related to factors which causing Celiac Disease marked with R and areas in gray are where the factors intersect in some parameter. Areas surrounded by bold frames are final disease causing physiological parameters which are part of matrix Pm

| Physiological Parameters related to external factors<br>(1) | External factors causing Celiac Disease |  |  |  |  |  |  |
| --- | --- | --- | --- | --- | --- | --- | --- |
| | 3+ respiratory infections in the first 6 months ( 1 cs) | Gluten before 2 years ( 1 cs) | Low IgA (2 cs) | HLA-DQ2/DQ8 common haplotypes ( 1cs) | Iron supplements at 18 months ( 1cs) | DDE, PFOS, perfluoro octanoic acid pesticides (2 cs) | TNF-α-308G>A polymorphism ( AA+AG vs GG) ( 1cs) |
| (1) | (2) | (3) | (4) | (5) | (6) | (7) | (8) |
| **Gliadin** |  | R |  |  |  |  |  |
| **glutenin** |  | R |  |  |  |  |  |
| **alfa,beta, gamma, omega-gliadin** |  |  |  |  |  |  |  |
| **Rothia bacteria** |  | R | R |  |  |  |  |
| **Staphylococcus epidermidis** |  | R | R |  |  |  |  |
| **Bifidobacterium** |  | R | R |  | R |  |  |
| **Alpha-gliadins 3 epitopes** |  | R |  |  |  |  |  |
| **Trypsin** |  | R |  |  |  |  |  |
| **Chymotrypsin** |  | R |  |  |  |  |  |
| **Carboxypeptidase A , B enzyme** |  | R |  |  |  |  |  |
| **Elastase** |  | R |  |  |  |  |  |
| **IFN-Gamma** | R | R |  |  |  |  |  |
| **IL-15** |  | R |  |  |  |  |  |
| **IgA** |  | R |  |  |  |  |  |
| **IgG** |  | R |  |  |  |  |  |
| **IFN Type 1** | R |  |  |  |  |  |  |
| **TFN -alfa increase** | R |  |  |  |  | R | R |
| CD4+ T-cell | R | R |  |  |  |  |  |
| Reovirus antibodies | R |  |  |  |  |  |  |
| IRF-1 | R |  |  |  |  |  |  |
| Reovirus T1L | R |  |  |  |  |  |  |
| IFN- beta | R |  |  |  |  |  |  |
| Microbiota |  |  | R |  |  |  |  |
| Proinflammatory IL-1beta, IL-6, TNF-alfa |  |  |  |  | R |  |  |
| IL-8 | R |  |  |  |  | R |  |
| IL-6 |  |  |  |  |  | R |  |

**Table 2.2.** Short list of physiological parmeters found to be causes of Celiac Disease and external factors impacting them.

| External Factors | Physiological parameters which changes are causing Celiac Disease |  |  |  |  |  |
| --- | --- | --- | --- | --- | --- | --- |
|  | Unknown physiological param 1 | Unknown physiological param 2 | Bifidobacterium, etc | IgA reduction | HLA-DQ2/DQ8 common haplotypes | TNF-alfa increase |
| 3+ respiratory infections in the first 6 months ( 1 cs) |  |  |  |  |  | X |
| Gluten before 2 years ( 1 cs) |  |  | X |  |  |  |
| Low IgA (2 cs) |  |  | X | X |  |  |
| Genetic predisposition ( 1 cs) |  |  |  |  | X |  |
| Iron supplements at 18 months ( 1cs) |  |  | X |  |  |  |
| DDE, PFOS, perfluorooctanoic acid pesticides (2 cs) |  |  |  |  |  | X |
| TNF-α -308G>A polymorphism ( AA+AG vs GG) ( 1cs) | | | | | | X |

This table is built using a Table 2.2 for factors / parameters for a Celiac disease by creating columns for physiological parameters changes causing the disease and placing the factors found from former table to the new one in the column with the physiological parameters in which the factor either causing a change or related to the change.

**Note:** the list of factors is obviously not complete and provided as per author’s knowledge of specific research, other factors can be added in the future if new research finds them, etc. At the same time the future factors as the current ones will be related to the physiological parameters in the columns of the table as these parameters the real causes of a disease (the factors are only causing their change).

For example, a research shows that “Enterovirus and adenovirus infections were diagnosed by seroconversions in virus antibodies in longitudinally collected sera using EIA. **Enterovirus infections were more frequent in case children before the appearance of celiac disease-associated tissue transglutaminase** autoantibodies compared to the corresponding period in control children (**OR 6.3**, 95% CI 1.8–22.3; p = 0.005) “(“Enterovirus Infections Are Associated With the Development of Celiac Disease in a Birth Cohort Study”, Front. Immunol., Maarit Oikarinen,Leena Puustinen,Jussi Lehtonen, et al, 02 February 2021, https://doi.org/10.3389/fimmu.2020.604529). As we can see **OR=6.3** and it means it is within a range of Disease Causing criteria for OR **4.55 +/- 50%** (2.27 - 6.81). So it means that *frequent Enterovirus infections in children is a disease causing factor* for Celiac disease and it is changing 1 physilogical parameter beyond 1-sigma. This parameter should be one of those 6 mentioned. As we know now, it cannot cause a Celiac disease standalone but together with other 5 factors it must trigger it. The appropriate physiological parameter need to be found using the preseneted method once more research is available.

## Results and Discussion

We introduced a method how to find a disease cause using a multiple researches which give estimates of risks (RR/OR) for different factors in regards to the *same* specific non-infectious disease. Method starts with finding a ***number of disease causes using a formula*** (1.0) when we know a rate of this non-infectious disease in a population (often we use an incidence rate for this to see annual probability of the disease).According to the method, once the risk factors were found using ***disease causation criteria provided above*** we need to find out physiological parameters which are changed when these factors are present.

Once we list all disease causing factors vertically and physiological parameters *related* to them horizontally (or vice versa) we mark the relationship between related factors and physiological parameters changed with a letter **“R”**. Then we find so called intersections between 2 factors - the places where 2 different factors are having same physiological parameters marked as related with letter “R”. The parameter are same by name (not by value).

**Table 2.3.**
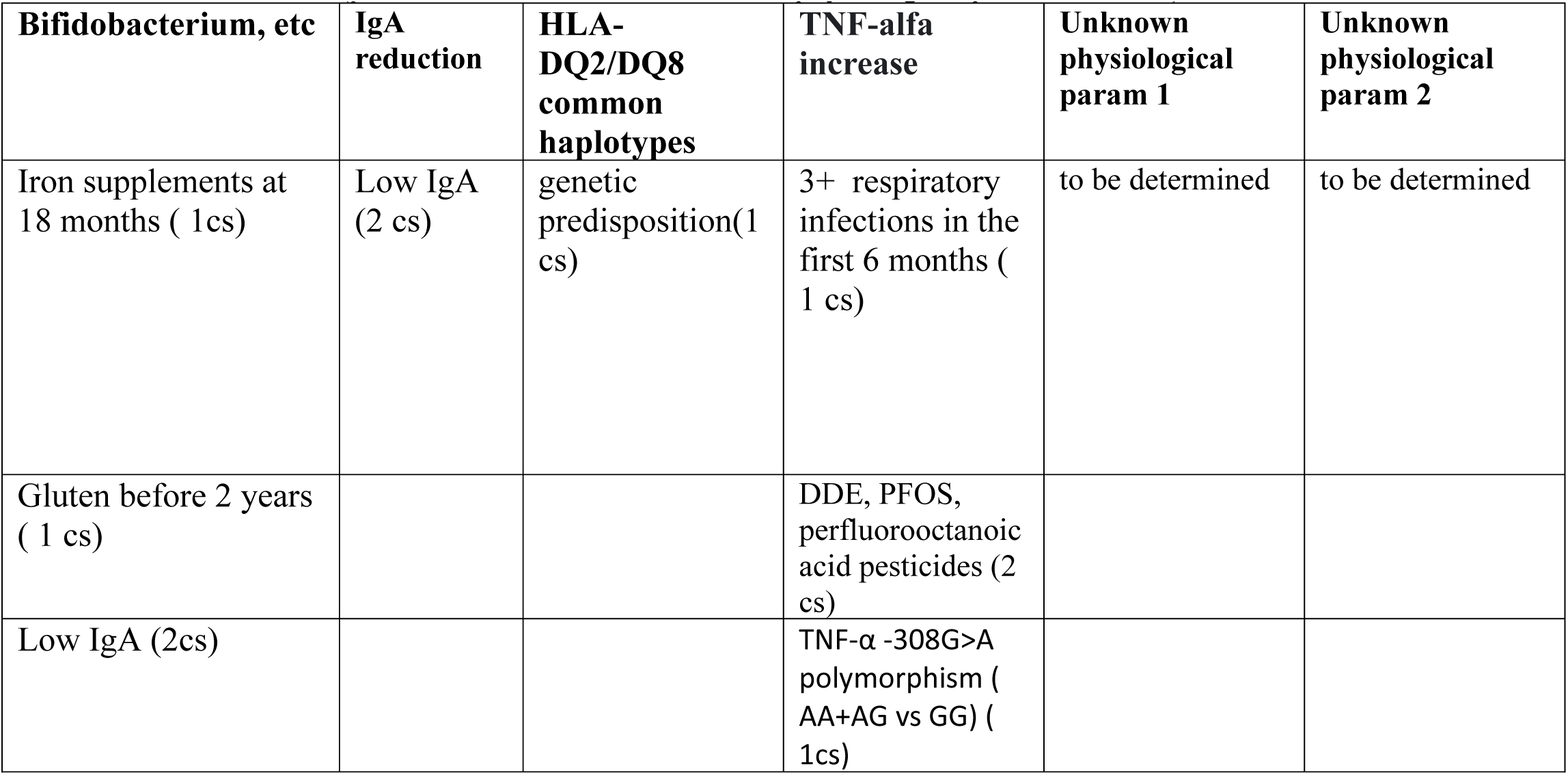
External factors which can cause the Celiac Disease. Any factor combination taken for all different columns should cause a Celiac disease (plus factors taken from 2 unknown physiological param columns)

| **Bifidobacterium, etc** | **IgA reduction** | **HLA-DQ2/DQ8 common haplotypes** | **TNF-alfa increase** | **Unknown physiological param 1** | **Unknown physiological param 2** |
| --- | --- | --- | --- | --- | --- |
| Iron supplements at 18 months ( 1cs) | Low IgA (2 cs) | genetic predisposition(1 cs) | 3+ respiratory infections in the first 6 months ( 1 cs) | to be determined | to be determined |
| Gluten before 2 years ( 1 cs) |  |  | DDE, PFOS, perfluorooctanoic acid pesticides (2 cs) |  |  |
| Low IgA (2cs) | | | TNF-α -308G>A polymorphism ( AA+AG vs GG) ( 1cs) | | |

We mark the column in gray or other color to indicate an intersection. We need to find all intersections like this. The physiological parameters where intersection are happening we list as matrix **Pm**. This list or matrix **Pm** is a superset of the physiological parameters changes which are causing a disease if their changes go beyond 1-sigma interval (actually, slightly less than this interval) and are present at the same time.

We eliminate the redundant parameters from this matrix **Pm** by using *few rules of elimination* and following a recommended algorithm if the count of parameters found exceeds the count determined by formula (1.0) calculating the number of disease causes or if a strict rule that a factor should cause only 1 change(or as determined by Disease Causation criteria) is broken.

One of these elimination rules is to check if the factors where intersection happens are related to each other (for example the same type of disease, or similar experiment like BMI and weight impact on disease, etc) and ignore the intersection for this factors as they are likely due to similarity. Another rule is to check if the physiological parameters for the same factors are related and this causing a change in 2 parameters for the same factor as one valid parameter is changing a dependent one. The dependent parameter need be eliminated from matrix **Pm**. Remember that most factors in practice are impacting only 1 physiological parameter change which is causing a disease. If we see a factor with 1 calculated impact is found to be causing 2 or more changes in physiological parameters in our table then the redundant physiological parameters need to be eliminated using the elimination rules.

Once the redundant parameters were eliminated we can check the final list of physiological parameters or matrix **Pm** using our *criteria for disease causing factors* (provided above for your reference) if there is an appropriate research regarding risks of these physiological parameters for the disease and confirm that we did not make any errors in steps of the described method. Also, we can use a related research regarding these found physiological parameters to see significance of their impact on the disease and this way to confirm the significance of the found physiological parameters found via the described method.

As example of method’s application we analyzed few non-infectious diseases. One was Depression and another was Celiac disease. We found that according to the method **Depression is caused by 2 physiological parameters changes** beyond 1-sigma interval if they present at the same time. The disease must happen if the disease causing condition is satisfied for some time. We notice that Depression should *not* happen if only 1 of these parameters is beyond 1-sigma interval. **The physiological parameters which must causes a Depression in an individual are Reduction of GABAergic inhibition beyond ∼1 sigma interval and decrease of BDNF beyond ∼1 sigma**. By controlling these 2 parameters we should be able to prevent Depression or possibly cure if the disease is in early stage.

In another example of method’s application we analyzed Celiac disease. We determined that **Celiac disease happens due to 6 physiological parameters changes** beyond 1-sigma interval according to the method and a mathematical model it is using. We determined that **the physiological parameters which are causing Celiac disease are:**

1. Bacteria Bifidobacterium, etc. changes in the intestine beyond ∼1 sigma
2. Reduction of immunologlobulin IgA beyond ∼1 sigma
3. Genetic predisposition - HLA-DQ2/DQ8 common haplotypes
4. TNF-alfa increase beyond ∼1 sigma
5. 2 physiological parameters changes beyond ∼1 sigma unknown and need to be determined

By controlling 4 of these causation factors (we cannot control genetics) we can prevent Celiac disease in an individual or possibly cure (if a disease in early stages) or treat it. If all the required physiological parameters changes take place at the same time the **Celiac disease *must*** start after some time unless they are taken under control by forcing them to be within 1-sigma interval. The **triggering of the disease if causation conditions are satisfied is not optional.** The analogy of this process could be an electrical fuse in a house. We know that if we plug in an electrical iron, air conditioner, etc. and exceed the threshold for the fuse the fuse will burn out. The fuse must burn out, it is not optional. The only random facts here are the electrical appliances which will cause it, these electrical appliances are an analogy of factors causing diseases here. We can have different external factors causing a disease but as long as they impact the all required physiological parameters a “fuse” of the body will burn out and the disease will start.

We need to notice here that multiple medical researches **often find risk factors for a specific disease which have values close to RR/OR = 4.6 or RR/OR = 20.6**. These are not just a statistically significant risks but these numbers indicate that 1) the **factors are really disease causing factors** 2) the factors are **causing a change in 1 physiological parameter beyond 1-sigma** (for RR/OR = 4.6) or **2 physiological parameters beyond 1-sigma** (for RR/OR = 20.6). Examples of these cases were shown in this article for gluten or pesticides researches for Celiac disease. In some rare cases the risk factors can have **RR/OR=93**+/-50% and this means the disease causing factors are impacting **3 physiological parameters beyond 1-sigma.** The disease causation criteria provided in article *“A Connection between Factors Causing Diseases and Diseases Frequencies: Its Application in Finding Disease Causes***”** (*Alan Olan,* Journal of Clinical Trials, Vol.13, Issue 4) allows to detemine more precisely if the risk factors are causing a disease or not and allows to expand validity for these risks beyond the 2 examples of RR/OR above. The referred article is using a risk factor **K** but it is easily converted for RR/OR as **RR = K + 1** (same for OR) and the RR/OR can be used as the theoretical values for RR/OR +/-50%. A short reference of these criteria was provided in this article as well.

In this article we have shown that **disease causation factors can be separated into few groups** according to a physiological parameter they change beyond 1-sigma. **Any combination of factors** taken as one from each of these groups **must cause a non-infectious disease** (one example is Table 2.3 for Celiac disease). For example “Iron Supplementation at 18 month”(1cs) factor AND “Low IgA”(2cs) AND “TNF alfa AA+AG polymorphism”(1cs) + “HLA-DQ2/DQ8 common haplotypes”(1cs) AND presence of 2 unknown factors (2cs) taken to make a sum of **6** found causes for Celiac disease *must* trigger the Celiac disease after some time. Please note that the factors are having a symbolic name and for their precise meaning refer to the research citation provided in this article.

We also showed how a knowledge of disease causes (as physiological parameters changed beyond 1 sigma) allows to build a hypothesis of disease pathology more easy as we know a disease “structure” and just need to “connect the dots” of it.

In Appendix we introduce **a principle of indifference** which we find as a useful tool in understanding of disease causes as well.

## Conclusions

In this article we have showed how a research which estimate a risk of non-infectious disease can be used to determine disease cause as a set of physiological parameters changes beyond 1-sigma (actually, slightly less than this interval). We showed how to determine which physiological parameter is impacted by a specific external factors. It was shown how certain different combinations of external factors can cause a disease if they act together and how to determine these disease causing factors combinations.

Overall a method presented here allows to increase efficiency of existing medical research in finding non-infectious disease causes and also stresses that non-infectious disease are having a multi-factorial causes. These causes are multiple physiological parameters changes beyond 1-sigma occurring at the same time and happening due to actions of external disease causing factors.

by Alan Olan - 2023

**Disclaimer:** This work is not intended to be used in diagnosing and treatment of any disease in any patient, those actions should be done by qualified professionals only. The work is intended as a reference for the researchers only. The author is not responsible for any issues created by unintentional usage or consequences of this work.

## Data Availability

All data produced in the present work are contained in the manuscript

## APPENDIX

### 1. Analogy to clarify a method

A **simplified analogy** which explains why we looking for a match between physiological parameters could be this. Imagine a large town. We observe it from the top. A person arrives to downtown for some *personal business* on a regular day. How likely this person meet *a friend* or colleague in a totally random place of the downtown?..Very unlikely. We observe this person is going to downtown for few days to different places and he never meets a friend or a colleague. Now, we observe from the top that some rare folks meet someone in downtown often and sometime in the same location. We know **there is a pattern explaining these meetings**, they are not random in most cases. There might be someone they have agreed to meet with before (a colleague they travel together with, a friend, etc). If we find these folks meeting we know we found very likely some pattern.

In this analogy, we can treat a downtown as a human body, a person arriving to downtown as a physiological parameter change caused by some disease causing factor. People which meet each other in the downtown are an analogy of physiological parameters which “meet up” as they cause a disease and not just a random meeting. If we find those folks who meet up we know there is some cause there. In the method presented, these “meetings” between physiological parameters are represented by “intersections”.

### 2. Principle of Indifference

As we discuss a method to find disease causes via experiments we are proposing to use a simple principle which can be observed in physiological processes. **When regulating its own homeostasis separate physiological systems of the body are indifferent to the side effects of this regulation.** We can call this *a principle of indifference*.

We can observe this principle in multiple cases. For example, if a brain needs to increase a blood pressure to improve a supply of nutrients and oxygen it is *indifferent* to the fact that a heart may not be able to sustain this high blood pressure. Another example could be this. If an intestine is trying to regulate its homeostatis of bacteria it can pass the signals to the brain via different biochemical pathways to reduce appetite so in this case an intestine is *indifferent* to that its actions can eliminate some source of energy and needed nutrients to the brain and other sysems and worsen their functioning.

This principle allows to explain that supporting a homeostasis in one physiological system of the body can and often does harm functioning of other systems and this way it can cause a non-infectious diseases.If we find a physiological system which is regulating its homeostasis with extreme steps and help it to fix the problems we can address a non-infectious disease of another physiological system which can be caused by this indifference.

